# SARS-CoV-2 serology in 4000 health care and administrative staff across seven sites in Lombardy, Italy

**DOI:** 10.1101/2020.05.24.20111245

**Authors:** Maria Teresa Sandri, Elena Azzolini, Valter Torri, Sara Carloni, Chiara Pozzi, Michela Salvatici, Michele Tedeschi, Massimo Castoldi, Alberto Mantovani, Maria Rescigno

## Abstract

Lombardy is the Italian region most affected by COVID-19. We tested the presence of plasma anti-SARS-CoV-2 IgG antibodies in 3985 employees across 7 healthcare facilities in areas of Lombardy with different exposure to the SARS-CoV-2 epidemic. Subjects filled a questionnaire to self-report on COVID-19 symptoms, co-morbidities, smoking, regular or smart-working, and the exposure to COVID-infected individuals. We show that the number of individuals exposed to the virus depended on the geographical location of the facility, ranging between 3 and 43%, consistent with the spatial variation of COVID-19 incidence in Lombardy, and correlated with family contacts. We observed a higher prevalence of females than males positive for IgG, however the level of antibodies was similar, suggesting a comparable magnitude of the response. IgG positivity among smokers was lower (7.4% vs 13.5%) although without difference in IgG plasma levels. We observed 11.9% of IgG positive asymptomatic individuals and another 23.1% with one or two symptoms. Interestingly, among the IgG positive population, 81.2% of subjects with anosmia/dysgeusia and fever were SARS-CoV-2 infected, indicating that these symptoms are strongly associated to COVID-19. The plasma level of IgG inversely correlated with anti-pneumococcal vaccination.

In conclusion, the frequency of IgG positivity and SARS-CoV-2 infection is dependent on the geographical exposure to the virus and primarily to family rather than hospital exposure.

## Introduction

Lombardy has been the Italian region most affected by SARS-Cov-2 infection with more than 80,000 cases from the outbreak and a mortality rate of nearly 18% (as of May 8th, https://utils.cedsdigital.it/coronavirus/#regioni1). Only symptomatic individuals have been tested by RT-PCR to detect SARS-CoV-2, and hence the spreading of the virus, particularly in the asymptomatic population, is unknown. Detection of viral RNA is not very sensitive in cases of low viral load, when patients with negative RT PCR test convert to positive in few days, or when COVID-19 is highly suspected by the result of chest CT ^1-6^. It is just a snapshot of the exact moment of the infection. Hence, unless being continuously monitored by viral testing, the asymptomatic individuals’ infected state may be missed.

A retrospective way to assess viral spread is via the analysis of an immune response that can be detected through the development of immunoglobulins ^7^. Indeed, almost the totality (>95%) of laboratory-confirmed COVID-19 showed seroconversion even using different methodologies and antibody isotypes (ELISA, EIA, anti-IgM, anti-IgG or anti-IgA) ^7-11^. Interestingly, the dynamic of the immune response to SARS-CoV-2 is very peculiar and patient dependent. IgM for instance, which should be the first type of immunoglobulins to appear under normal circumstances, are detected before, concomitant or even after IgGs, or do not appear at all ^7,12^. This suggests that differently from other infectious diseases, in SARS-CoV-2 infection, IgG is a more reliable marker for seroconversion than IgM.

There is evidence that IgG analysis allowed to detect viral RNA negative asymptomatic relatives of COVID-19 patients ^7^, suggesting that anti-SARS-CoV-2 IgG may be used as a diagnostic tool to detect the incidence of asymptomatic infections even in cases of impossibility of viral detection. Based on these assumptions, we tested the presence of plasma anti-SARS-CoV-2 IgG antibodies in nearly 4000 (3985) employees of 7 different healthcare facilities, one including a research center and a University, located across the Lombardy region in areas with different exposure to the epidemic (Milan, Rozzano, Varese, Castellanza and Bergamo (the city most affected by the pandemic per number of individuals)). All of the analyzed hospitals drastically reduced their routine practice and, except for Humanitas Medical Care, which is a point of care, they were all dedicated to COVID-19 during the time of observation.

## Results

### Study population

Recruitment was on a voluntary basis: it started on April 28^th^ and more than 65% of employees participated as of May 16^th^, 2020 (clinicaltrial.gov NCT04387929). An international medical school (Humanitas University, Pieve Emanuele (MI)) and 6 different hospitals participated to the study: Istituto Clinico Humanitas (ICH), Rozzano (MI); Humanitas Gavazzeni, Bergamo; Humanitas Castelli, Bergamo; Humanitas Mater Domini (HMD), Castellanza (VA); Humanitas Medical Care (HMC), Varese (VA); Humanitas San Pio X, Milano (MI). The individuals were healthcare professionals (physicians (16.5%), surgeons (7.2%) anesthesiologists (2.8%), physiotherapists (1.8%), nurses (25.4%)), technicians (4.4%), students (0.7%), researchers/academics (3.1%), other roles, including administrative staff (7.9%), biologists (1.3%) etc.. 66.8% were females and 33.2% males, median age 42 yo (21-86 yo) (Supplementary Table S1). The distribution of the healthcare workers participating to the study among the different hospitals was very similar except for HMC, a point of care, where the proportion of workers differs from that of the other hospitals, and for ICH which is the only site where researchers work (Supplementary Fig. S1). Besides this, we did not observe major differences in the voluntary participation of the different professionals across sites.

Subjects filled a questionnaire reporting whether they had COVID-associated symptoms, including fever, dry cough, asthenia, soreness and muscle pain, runny nose, sore throat, gastrointestinal symptoms (including vomiting, nausea, diarrhea), conjunctivitis, pneumonia, severe acute respiratory syndrome, anosmia/dysgeusia from February 1^st^ to the time of questionnaire filling. They also reported on body mass index (BMI), co-morbidities (hypertension, heart disease or diabetes, immunosuppression), smoking, vaccinations, regular or smart-working, the frequency of home exits for non-work related reasons and the exposure to COVID-infected individuals. With the exception of HMC, which being a point of care closed down at lockdown (officially started on March 9^th^), during the pandemic and in the period of observation, most of routine practice was interrupted, and the majority of physician and non-physician healthcare professionals worked in COVID-dedicated areas independent on the location of the hospital (for instance at ICH, 8 out of 20 wards were transformed into COVID treatment wards (330 beds), and 2 into semi-intensive care units, while the intensive care units (ICU) capacity doubled (60 beds)). All of the personnel working in the emergency room or customer care had to wear obligatory PPE from February 1^st^ at Castelli and Gavazzeni, from February 23^rd^ at ICH, and from March 3^rd^ everywhere in the hospital. Administrative staff workers stopped working in the hospital at the beginning of lockdown and worked from home. Customer service and research personnel worked one or two days a week at the sites and the rest of the time from home.

### Frequency of IgG positivity and correlation with geographical area

Anti-S1 and anti-S2 SARS-CoV-2 IgG specific antibodies were detected via an indirect chemiluminescence immunoassay. According to kit manufacturer, the test discriminates among negative (<12AU/mL; with 3.8 as the limit of IgG detection), equivocal (12.0 – 15.0 AU/mL) and truly positive (>15.0 AU/mL) subjects. Of 3985 enrolled subjects, 3462 (87%) were negative, 76 (2%) equivocal and 447 (11%) positive (which together with equivocal summed up to 523 (13%)) (Supplementary Table S1). We decided to group together equivocal and truly positive subjects as they behaved very similarly both as separate groups or as grouped together (Positive - IgG ≥ 12.0 AU/ml) in subsequent analyses (for instance in correlation with the number of symptoms, as shown in Supplementary Fig. S2).

The number of individuals exposed to the virus reflected the geographical area where the healthcare facility was located (Fig. 1). Two hospitals based in Bergamo (Humanitas Gavazzeni and Castelli) were the most affected as between 35 and 43 % of the subjects was IgG positive. In the other sites the frequency of IgG positivity ranged between 3% (HMC) and 9% (ICH) (Supplementary Table S2). We applied a hierarchical (multilevel) model adjusting for clustering of individuals within healthcare facilities when assessing overall relationships between locations, professional status and the analyzed variables. Humanitas Gavazzeni in Bergamo was significantly different from all other facilities except for Castelli in Bergamo (Table 1 and Supplementary Fig. S3, *p*<0.0001,OR and 95%CI indicated in the table). Interestingly, HMC and HMD which had the lowest incidence of IgG positive individuals (3% and 3.8%, respectively), are located in Varese, which is one of the less COVID-19 affected province in Lombardy (https://www.lombardianotizie.online/coronavirus-casi-lombardia/). In agreement, the percentage of patients hospitalized for COVID-19 was higher in Bergamo at Gavazzeni (56%), intermediate in Milan at ICH (36%) and low in Varese at HMD (10%) (not shown).

**Table 1.** Logistic analysis for site effect

| Table 1 Logistic analysis for site effect |  |  |  |  |
| --- | --- | --- | --- | --- |
| Comparison | Odds ratio estimates | Lower 95% CI | Upper 95% CI | p-value |
| Humanitas Rozzano (ICH) | 0.181 | 0.146 | 0.226 | <.0001 |
| Humanitas University (HU) | 0.156 | 0.062 | 0.396 |  |
| Humanitas Medical Care (HMC) | 0.057 | 0.014 | 0.234 |  |
| Humanitas San Pio X | 0.126 | 0.074 | 0.215 |  |
| Humanitas Mater Domini (HMD) | 0.073 | 0.041 | 0.131 |  |
| Humanitas Castelli | 1.384 | 0.943 | 2.032 |  |
| Humanitas Gavazzeni - ref | 1.00 |  |  |  |

**Figure 1.**
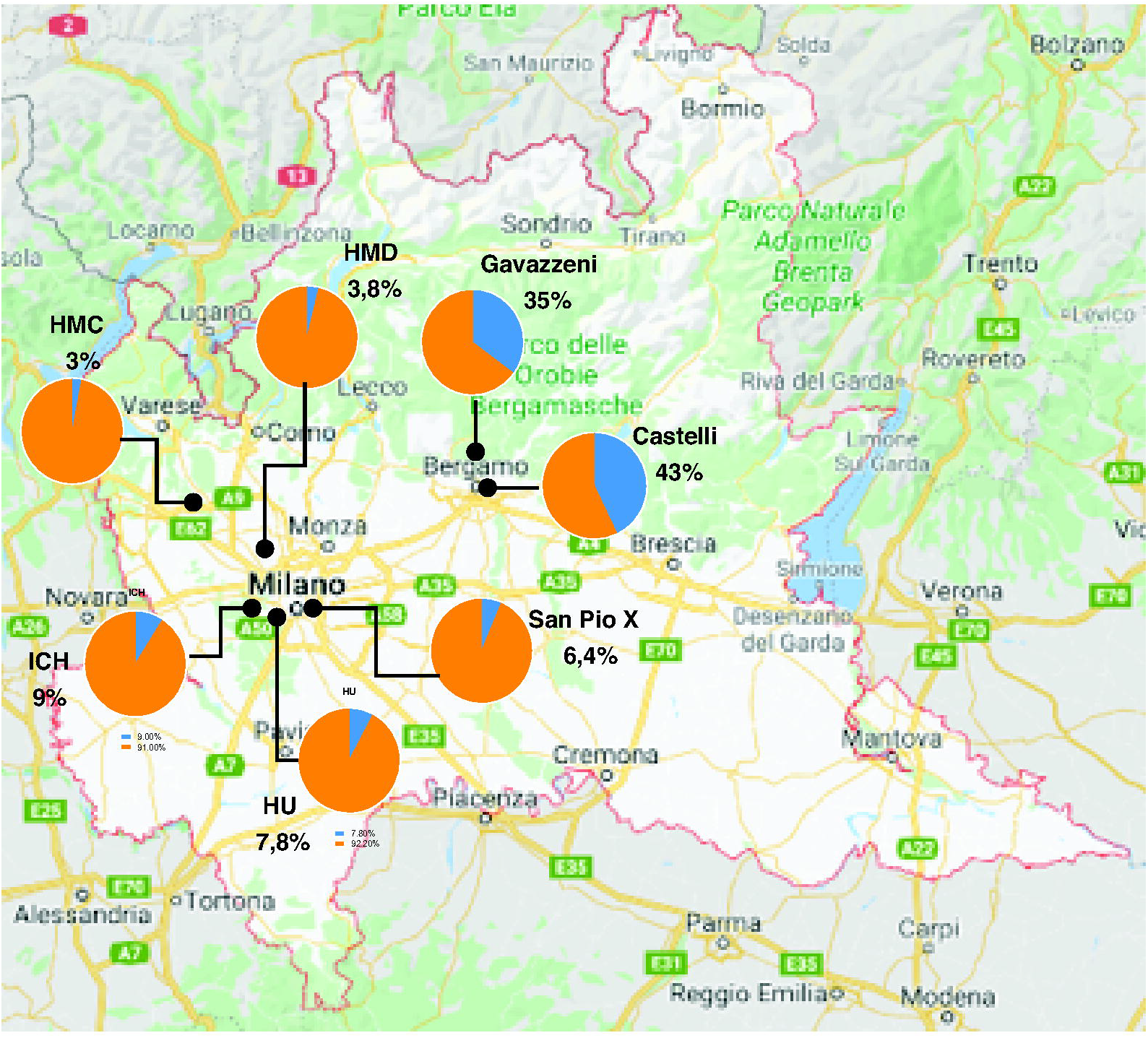
Distribution of IgG positivity (≥12 AU/mL) across different geographical areas and healthcare facilities in Lombardy, Italy. Employees from 7 different healthcare facilities were analyzed for their anti-SARS-CoV-2 IgG positivity. Pie charts show the percentage of negative subjects (IgG <12 AU/mL) (orange) and that of positive subjects (IgG ≥12 AU/mL) (blue) in each site: Istituto Clinico Humanitas (ICH), Rozzano (MI); Humanitas Gavazzeni, Bergamo; Humanitas Castelli, Bergamo; Humanitas Mater Domini (HMD), Castellanza (VA); Humanitas Medical Center (HMC), Varese (VA); Humanitas University (HU), Pieve Emanuele (MI); Humanitas San Pio X, Milan, (MI).

### Frequency of IgG positivity and correlation with gender, BMI and smoking

IgG positivity (≥12 AU/mL) was different between males (11.4%) and females (14%) (Supplementary Table S1), and, as shown in Table 2a, this difference was retained in the multilevel logistic analysis (OR=0.73; 95%CI 0.57-0.94, *p=*0.016). Thus, the observed effect was associated to a difference in gender sensitivity to infection or to the ability to mount an antibody response rather than to a bias of the recruitment itself.

**Table 2a.** Multilevel Logistic model for age, gender and their interaction, smoking habits, BMI

|  |  |  |  |  |
| --- | --- | --- | --- | --- |
| Comparison | Odds ratio estimates | Lower 95% CI | Upper 95% CI | p-value |
| Physician vs Other Role | 1.10 | 0.79 | 1.53 | 0.5039 |
| Surgeon vs Other Role | 1.10 | 0.71 | 1.71 |  |
| Anesthesiologist vs Other Role | 0.49 | 0.22 | 1.08 |  |
| Physiotherapist vs Other Role | 1.17 | 0.56 | 2.46 |  |
| Nurse vs Other Role | 1.16 | 0.87 | 1.55 |  |
| Researcher vs Other Role | 0.94 | 0.47 | 1.90 |  |
| Biologist vs Other Role | 0.69 | 0.20 | 2.37 |  |
| Student vs Other Role | 1.07 | 0.29 | 3.95 |  |
| Radiology Technician vs Other Role | 0.53 | 0.25 | 1.13 |  |
| Lab. Technician vs Other Role | 0.75 | 0.31 | 1.86 |  |
| Staff vs Other Role | 1.04 | 0.67 | 1.61 |  |
| Smoking (Number of cigarettes/day) | 0.48 | 0.38 | 0.61 | <.0001 |
| Age >60 vs $\leq 60$ | 0.35 | 0.15 | 0.83 | 0.0166 |
| Gender male vs female | 0.73 | 0.57 | 0.94 | 0.0158 |
| Interaction Age*Gender | 2.98 | 1.08 | 8.21 | 0.0352 |
| BMI | 1.11 | 0.98 | 1.26 | 0.1127 |

**Table 2b.** Multilevel Logistic model for symptoms adjusted for professional status, age, gender and their interaction, smoking habits, BMI

|  |  |  |  |  |
| --- | --- | --- | --- | --- |
| Comparison | Odds ratio estimates | Lower 95% CI | Upper 95% CI | p-value |
| Fever | 5.79 | 3.58 | 9.36 | <.0001 |
| Low-grade Fever | 1.41 | 0.86 | 2.33 | 0.1735 |
| Cough | 1.08 | 0.69 | 1.71 | 0.7266 |
| Sore Throat/Runny nose | 0.93 | 0.61 | 1.42 | 0.7332 |
| Muscle pain | 1.31 | 0.83 | 2.06 | 0.2453 |
| Asthenia | 0.92 | 0.57 | 1.47 | 0.7263 |
| Anosmia/Dysgeusia | 11.51 | 7.06 | 18.77 | <.0001 |
| Gastrointestinal disorders | 0.77 | 0.49 | 1.18 | 0.2299 |
| Conjunctivitis | 1.35 | 0.77 | 2.36 | 0.2887 |
| Dyspnea | 1.14 | 0.64 | 2.06 | 0.6539 |
| Chest pain | 2.17 | 1.26 | 3.74 | 0.0055 |
| Tachycardia | 0.64 | 0.38 | 1.09 | 0.0999 |
| Pneumonia | 9.00 | 1.34 | 60.66 | 0.0240 |

Interestingly, IgG positivity among smokers was lower (7.4% vs 13.5%) (OR=0.45; 95%CI 0.34-0.60, *p*<0.0001, Supplementary Fig. S4) confirming the trend effect observed with the multilevel logistic analysis (OR=0.48; 95%CI 0.38-0.61, *p*<0.0001, Table 2a). On the contrary, there was no statistically significant difference according to BMI (Table 2a).

### Frequency of IgG positivity and correlation with age

We then evaluated whether there was a difference in the positivity to IgG according to age. We found that there was a Gaussian distribution of the number of IgG positive (≥12 AU/mL) individuals across the age range (Fig. 2a), but then when analyzing the frequency of positivity at the different age ranges we observed an age dependent reduction of IgG positive individuals (Fig. 2b). However, this age-dependency was primarily due to the female rather than the male population, particularly for subjects either young (20-40 yo) or older than 60 yo (test for heterogeneity of trend effect age between male and female *p*=0.0024 Fig. 2b). In older than 60 yo, IgG positivity dropped from 12% in males to 5% in females (Table 2a and Fig. 2b). This indicates that females are more likely to be infected – or to induce an IgG response - when young, and less likely at ages higher than 60 yo.

**Figure 2.**
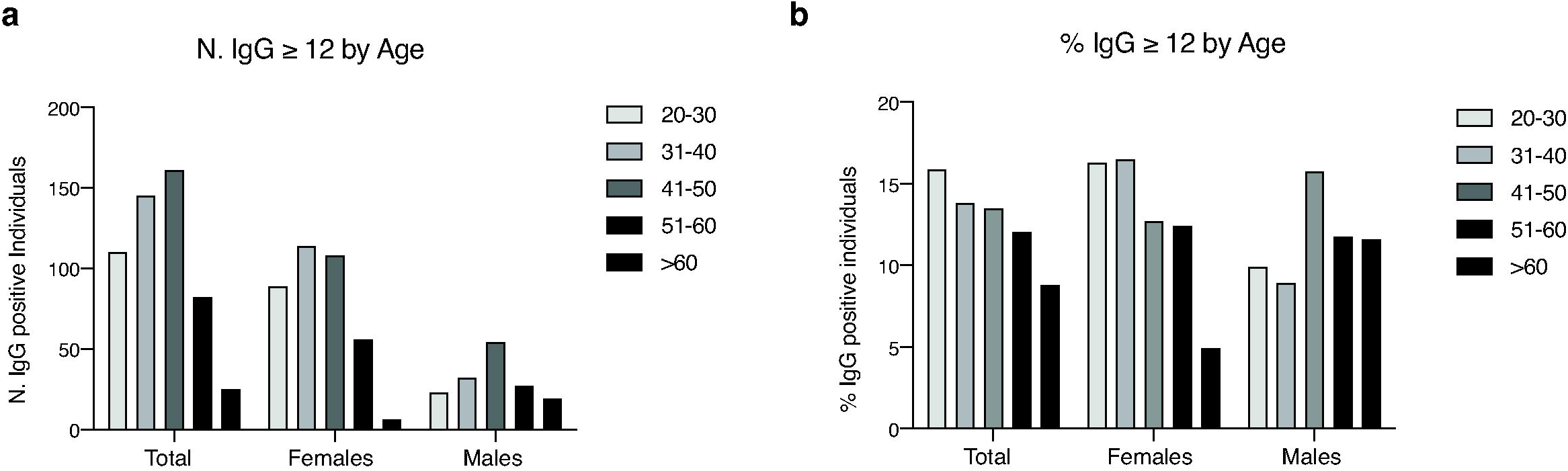
Frequency of IgG positivity (IgG ≥ 12 AU/mL) by age. a, b, Histograms show the number (a) and the percentage (b) of positive individuals (IgG ≥ 12 AU/mL) divided by age range and sex on the whole population regardless of site; in b, *p*-values were determined using Cuzick’s test for trend. *p* = 0.0374 (total); *p* = 0.0008 (female); *p* = 0.1498 (male); *p* = 0.0024 (interaction).

### Frequency of IgG positivity and correlation with viral positivity at time of testing

Following the guidelines of the national health system, all 523 subjects with IgG ≥ 12 AU/ml underwent a rinopharyngeal swab for SARS-CoV-2 RNA viral detection. We found that 39 (7.6%) individuals resulted positive for viral RNA detection (Table 3). However, in 31 of these (79.5%), the subjects tested negative for at least one of the genomic sequences of the three SARS-CoV-2 gene targets: *E, RdRp* and *N*. To rule out that the negative population (3.8<IgG<12 AU/mL) comprised individuals in the early phases of viral infection, a sample of 283 (35.4%) individuals underwent a rinopharyngeal swab for SARS-CoV-2 RNA viral detection. All of them resulted negative to the swab confirming the negativity of the test below 12 AU/mL (Table 3).

**Table 3.** Rinopharyngeal swab positivity in relation to IgG positivity

|  | Rinopharyngeal swab results |  |  |  |  |  |  |  |  |  |  |  |  |  |
| --- | --- | --- | --- | --- | --- | --- | --- | --- | --- | --- | --- | --- | --- | --- |
|  | Total |  | Negative |  |  | Positive low load |  |  | Positive |  |  | Positive low load + Positive |  |  |
|  | N | % of total | N | % of level | % of total | N | % of level | % of total | N | % of level | % of total | N | % of level | % of total |
| **IgG results** |  |  |  |  |  |  |  |  |  |  |  |  |  |  |
| Negative (3.8<IgG<12AU/mL) | 283 | 35.4 | 283 | 100.0 | 37.2 | . | . | . | . | . | . | . | . | . |
| Equivocal (12÷15AU/mL) | 75 | 9.4 | 70 | 93.3 | 9.2 | 3 | 4.0 | 9.7 | 2 | 2.7 | 25.0 | 5 | 6.7 | 12.8 |
| Truly Positive (>15AU/mL) | 441 | 55.2 | 407 | 92.3 | 53.6 | 28 | 6.3 | 90.3 | 6 | 1.4 | 75.0 | 34 | 7.7 | 87.2 |
| Positive (≥12AU/mL) | 516 | 64.6 | 477 | 92.4 | 62.8 | 31 | 6.0 | 100.0 | 8 | 1.6 | 100.0 | 39 | 7.6 | 100.0 |
| **Total** | 799 | 100.0 | 760 | 95.1 | 100.0 | 31 | 3.9 | 100.0 | 8 | 1.0 | 100.0 | 39 | 4.9 | 100.0 |

### Frequency of IgG positivity and correlation with professional status

We then evaluated the proportion of IgG positive individuals (≥ 12 AU/mL) across the different professional workers and found that there was no statistically significant difference when taking into account age, gender and their interaction, smoking habits, BMI and location (Table 2a) even though some professionals worked from home from the beginning of lockdown (such as the administrative staff), suggesting that their exposure to the virus was not occurring at work. We thus assessed which could have been the major driver of transmission of the virus on the basis of the self-reported questionnaire. We found heterogeneity of IgG positivity among drivers (*p*<0.0001). Compared with group referring no contact, IgG positivity correlated most with family contacts (OR=4.73; 95%CI 2.93-7.65), and not with either a colleague or a patient, suggesting that this was likely the major cause of viral exposure (Tables 4a-b). This finding is in line with a recent report on testing the serology of healthcare workers and their families ^13^.

**Table 4a.**
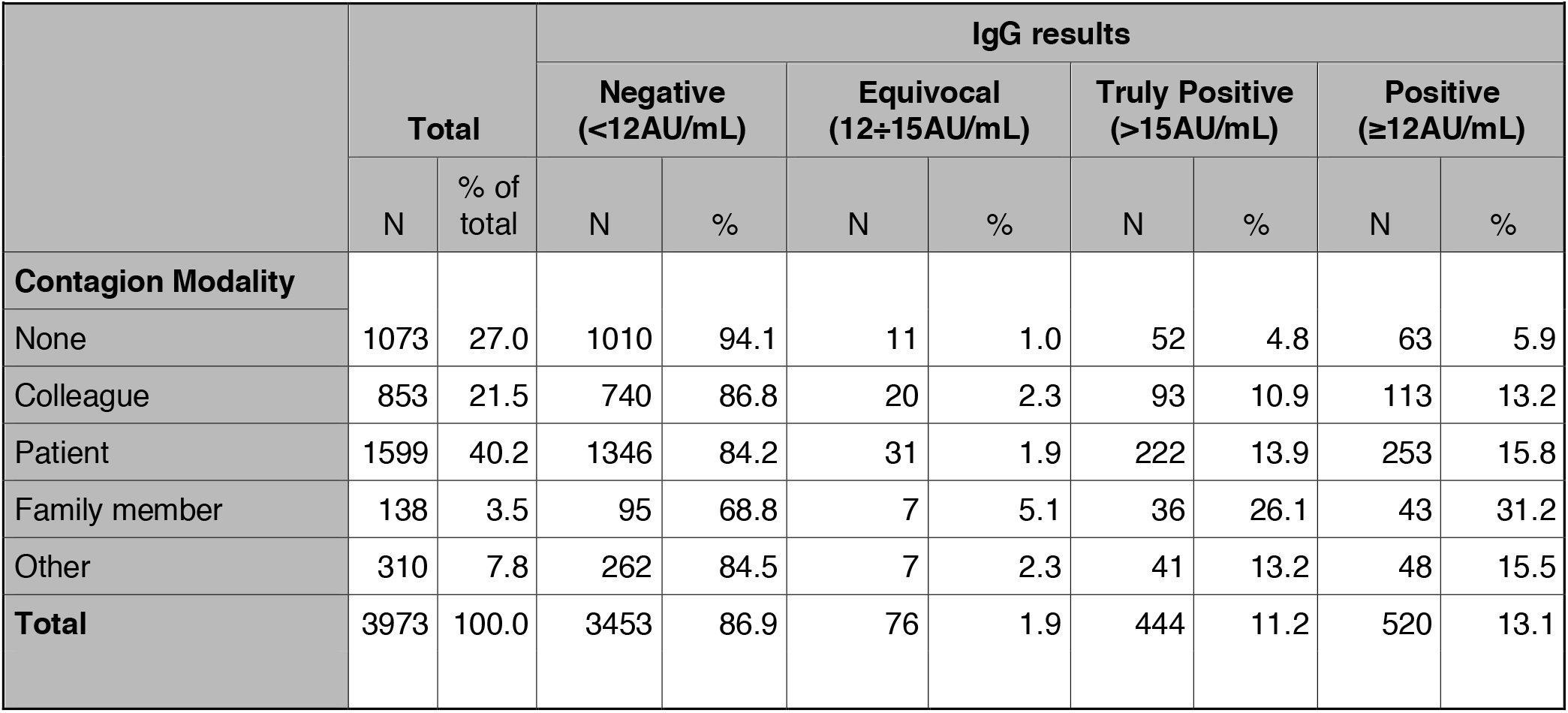
Association of contacts with IgG positivity

**Table 4b.** Summary measures of association of contacts with IgG positivity

|  |  |  |  |
| --- | --- | --- | --- |
| Contagion Modality | Odds ratio | 95% CI |  |
| None | Ref |  |  |
| Colleague | 1.59 | 1.11 | 2.27 |
| Patient | 1.83 | 1.29 | 2.60 |
| Family member | 4.73 | 2.93 | 7.65 |
| Other | 2.05 | 1.32 | 3.18 |
| P value (Wald test) | <0.0001 |  |  |
| Multilevel logistic analysis, considering subjects nested in the hospital site. Adjusted for role, age (cut off=60 years) gender, BMI, smoking habits. |  |  |  |

### Frequency of IgG positivity and correlation with symptoms, comorbidities and vaccinations

We then evaluated whether there was a correlation between the number and typology of self-reported symptoms and the frequency of antibody response both as number of individuals (Fig. 3a and 3b) and as percentage on the whole population (Fig. 3c and 3d). As shown in Figure 3, when analyzing individuals deemed to be positive on the basis of the amount of IgG (≥12 AU/mL) we observed a two phases decay: first a similar frequency (a) or number (c) of subjects with 0 to 7 concomitant symptoms, and then a drop of individuals at higher number of concomitant symptoms. This distribution followed a sigmoidal, four parameter logistic curve whereby X is the number of symptoms (R^2^=0.97). By contrast, in the population with IgG <12 AU/mL we found a higher number (b) or frequency (d) of individuals with 0, 1 or 2 symptoms and the distribution was following an exponential curve (R^2^=0.9975). The multilevel logistic analysis showed that, besides pneumonia, among the symptoms, fever, anosmia/dysgeusia (loss of smell or taste) and chest pain were those that best characterized the IgG positive population, particularly when collated (Table 2b).

**Figure 3.**
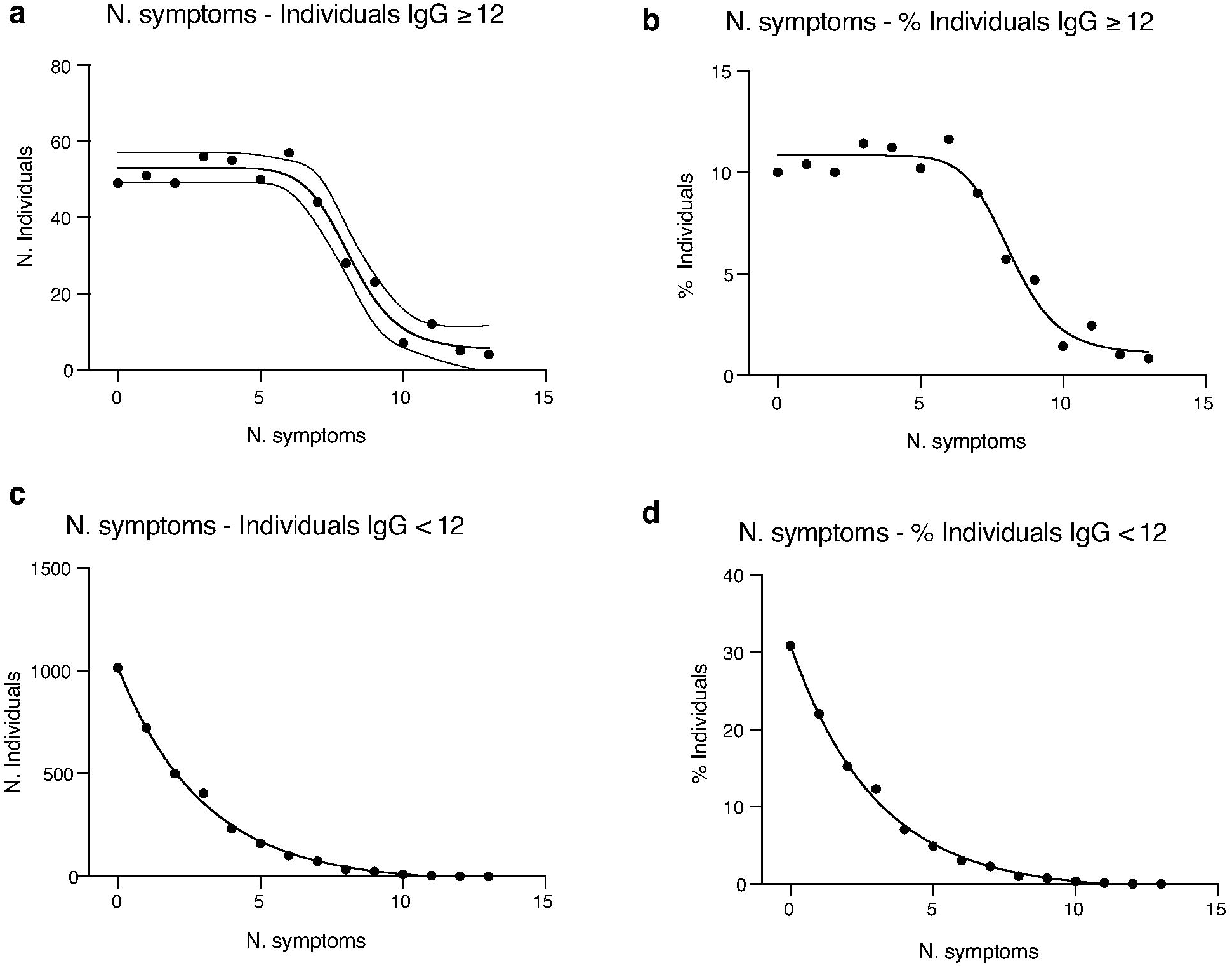
Distribution of the IgG ≥12 and IgG<12 populations versus the number of symptoms. a, c, Distribution of the IgG positive individuals (IgG ≥ 12 AU/mL) as number of individuals (a) or percentage of the population (c) versus the number of self-reported symptoms. The curve that best interpolated the data was a sigmoidal, four parameter logistic curve whereby X is the number of symptoms (R^2^=0.97). b, d, Distribution of the IgG negative individuals (IgG < 12 AU/mL) as number of individuals (b) or percentage of the population (d) versus the number of self-reported symptoms. The curve that best interpolated the data was exponential (R^2^=0.9975).

Indeed, dividing the population according to the number of symptoms, IgG positivity increased from 11.9% in the absence of symptoms to 42.8% in the presence of 5 symptoms or more (Table 5), and the value of the AUC derived by the multivariable model was 79%, while the combination of just fever versus anosmia/dysgeusia had an AUC of 78% (Supplementary Fig. S5). 81.2% of individuals presenting both anosmia/dysgeusia and fever resulted SARS-CoV-2 infected. The sensitivity, specificity and positive Likelihood ratio of anosmia/dysgeusia and fever were 25.6%, 99.1% and 28.6, respectively (Supplementary Table S3).

**Table 5.** Association of symptoms with IgG positivity

| Table 5 Association of symptoms with IgG positivity |  |  |  |  |  |  |  |  |  |  |  |  |  |  |
| --- | --- | --- | --- | --- | --- | --- | --- | --- | --- | --- | --- | --- | --- | --- |
|  | Total |  | IgG results |  |  |  |  |  |  |  |  |  |  |  |
|  |  |  | Negative (<12AU/mL) |  |  | Equivocal (12÷15AU/mL) |  |  | Truly Positive (>15AU/mL) |  |  | Positive (≥12AU/mL) |  |  |
|  | N | % of total | N | % of level | % of total | N | % of level | % of total | N | % of level | % of total | N | % of level | % of total |
| Fever |  |  |  |  |  |  |  |  |  |  |  |  |  |  |
| No | 3488 | 87.5 | 3179 | 91.1 | 91.8 | 56 | 1.6 | 73.7 | 253 | 7.3 | 56.6 | 309 | 8.9 | 59.1 |
| Yes | 497 | 12.5 | 283 | 56.9 | 8.2 | 20 | 4.0 | 26.3 | 194 | 39.0 | 43.4 | 214 | 43.1 | 40.9 |
| Low-grade Fever |  |  |  |  |  |  |  |  |  |  |  |  |  |  |
| No | 3557 | 89.3 | 3149 | 88.5 | 91.0 | 63 | 1.8 | 82.9 | 345 | 9.7 | 77.2 | 408 | 11.5 | 78.0 |
| Yes | 428 | 10.7 | 313 | 73.1 | 9.0 | 13 | 3.0 | 17.1 | 102 | 23.8 | 22.8 | 115 | 26.9 | 22.0 |
| Cough |  |  |  |  |  |  |  |  |  |  |  |  |  |  |
| No | 3074 | 77.1 | 2750 | 89.5 | 79.4 | 57 | 1.9 | 75.0 | 267 | 8.7 | 59.7 | 324 | 10.5 | 62.0 |
| Yes | 911 | 22.9 | 712 | 78.2 | 20.6 | 19 | 2.1 | 25.0 | 180 | 19.8 | 40.3 | 199 | 21.8 | 38.0 |
| Sore Throat/Runny nose |  |  |  |  |  |  |  |  |  |  |  |  |  |  |
| No | 2687 | 67.4 | 2391 | 89.0 | 69.1 | 50 | 1.9 | 65.8 | 246 | 9.2 | 55.0 | 296 | 11.0 | 56.6 |
| Yes | 1298 | 32.6 | 1071 | 82.5 | 30.9 | 26 | 2.0 | 34.2 | 201 | 15.5 | 45.0 | 227 | 17.5 | 43.4 |
| Muscle pain |  |  |  |  |  |  |  |  |  |  |  |  |  |  |
| No | 2957 | 74.2 | 2707 | 91.5 | 78.2 | 42 | 1.4 | 55.3 | 208 | 7.0 | 46.5 | 250 | 8.5 | 47.8 |
| Yes | 1028 | 25.8 | 755 | 73.4 | 21.8 | 34 | 3.3 | 44.7 | 239 | 23.2 | 53.5 | 273 | 26.6 | 52.2 |
| Asthenia |  |  |  |  |  |  |  |  |  |  |  |  |  |  |
| No | 3237 | 81.2 | 2948 | 91.1 | 85.2 | 43 | 1.3 | 56.6 | 246 | 7.6 | 55.0 | 289 | 8.9 | 55.3 |
| Yes | 748 | 18.8 | 514 | 68.7 | 14.8 | 33 | 4.4 | 43.4 | 201 | 26.9 | 45.0 | 234 | 31.3 | 44.7 |

Table 5 Association of symptoms with IgG positivity
|  | IgG results |  |  |  |  |  |  |  |  |  |  |  |  |  |
| --- | --- | --- | --- | --- | --- | --- | --- | --- | --- | --- | --- | --- | --- | --- |
|  | Total |  | Negative (<12AU/mL) |  |  | Equivocal (12÷15AU/mL) |  |  | Truly Positive (>15AU/mL) |  |  | Positive (≥12AU/mL) |  |  |
|  | N | % of total | N | % of level | % of total | N | % of level | % of total | N | % of level | % of total | N | % of level | % of total |
| **Anosmia/Dysgeusia** |  |  |  |  |  |  |  |  |  |  |  |  |  |  |
| No | 3617 | 90.8 | 3348 | 92.6 | 96.7 | 46 | 1.3 | 60.5 | 223 | 6.2 | 49.9 | 269 | 7.4 | 51.4 |
| Yes | 368 | 9.2 | 114 | 31.0 | 3.3 | 30 | 8.2 | 39.5 | 224 | 60.9 | 50.1 | 254 | 69.0 | 48.6 |
| **Gastrointestinal disorders** |  |  |  |  |  |  |  |  |  |  |  |  |  |  |
| No | 3165 | 79.4 | 2812 | 88.8 | 81.2 | 52 | 1.6 | 68.4 | 301 | 9.5 | 67.3 | 353 | 11.2 | 67.5 |
| Yes | 820 | 20.6 | 650 | 79.3 | 18.8 | 24 | 2.9 | 31.6 | 146 | 17.8 | 32.7 | 170 | 20.7 | 32.5 |
| **Conjunctivitis** |  |  |  |  |  |  |  |  |  |  |  |  |  |  |
| No | 3583 | 89.9 | 3145 | 87.8 | 90.8 | 64 | 1.8 | 84.2 | 374 | 10.4 | 83.7 | 438 | 12.2 | 83.7 |
| Yes | 402 | 10.1 | 317 | 78.9 | 9.2 | 12 | 3.0 | 15.8 | 73 | 18.2 | 16.3 | 85 | 21.1 | 16.3 |
| **Dyspnea** |  |  |  |  |  |  |  |  |  |  |  |  |  |  |
| No | 3726 | 93.5 | 3291 | 88.3 | 95.1 | 67 | 1.8 | 88.2 | 368 | 9.9 | 82.3 | 435 | 11.7 | 83.2 |
| Yes | 259 | 6.5 | 171 | 66.0 | 4.9 | 9 | 3.5 | 11.8 | 79 | 30.5 | 17.7 | 88 | 34.0 | 16.8 |
| **Chest pain** |  |  |  |  |  |  |  |  |  |  |  |  |  |  |
| No | 3664 | 91.9 | 3235 | 88.3 | 93.4 | 62 | 1.7 | 81.6 | 367 | 10.0 | 82.1 | 429 | 11.7 | 82.0 |
| Yes | 321 | 8.1 | 227 | 70.7 | 6.6 | 14 | 4.4 | 18.4 | 80 | 24.9 | 17.9 | 94 | 29.3 | 18.0 |
| **Tachycardia** |  |  |  |  |  |  |  |  |  |  |  |  |  |  |
| No | 3553 | 89.2 | 3112 | 87.6 | 89.9 | 62 | 1.7 | 81.6 | 379 | 10.7 | 84.8 | 441 | 12.4 | 84.3 |
| Yes | 432 | 10.8 | 350 | 81.0 | 10.1 | 14 | 3.2 | 18.4 | 68 | 15.7 | 15.2 | 82 | 19.0 | 15.7 |
| **Pneumonia** |  |  |  |  |  |  |  |  |  |  |  |  |  |  |
| No | 3951 | 99.1 | 3457 | 87.5 | 99.9 | 75 | 1.9 | 98.7 | 419 | 10.6 | 93.7 | 494 | 12.5 | 94.5 |
| Yes | 34 | 0.9 | 5 | 14.7 | 0.1 | 1 | 2.9 | 1.3 | 28 | 82.4 | 6.3 | 29 | 85.3 | 5.5 |
| **Other symptoms** |  |  |  |  |  |  |  |  |  |  |  |  |  |  |
| No | 3828 | 96.1 | 3341 | 87.3 | 96.5 | 71 | 1.9 | 93.4 | 416 | 10.9 | 93.1 | 487 | 12.7 | 93.1 |
| Yes | 157 | 3.9 | 121 | 77.1 | 3.5 | 5 | 3.2 | 6.6 | 31 | 19.7 | 6.9 | 36 | 22.9 | 6.9 |
| **Number of symptoms** |  |  |  |  |  |  |  |  |  |  |  |  |  |  |
| 0 symptoms | 1380 | 34.6 | 1318 | 95.5 | 38.1 | 11 | 0.8 | 14.5 | 51 | 3.7 | 11.4 | 62 | 4.5 | 11.9 |
| 1 symptoms | 814 | 20.4 | 758 | 93.1 | 21.9 | 10 | 1.2 | 13.2 | 46 | 5.7 | 10.3 | 56 | 6.9 | 10.7 |
| 2 symptoms | 582 | 14.6 | 517 | 88.8 | 14.9 | 11 | 1.9 | 14.5 | 54 | 9.3 | 12.1 | 65 | 11.2 | 12.4 |
| 3 symptoms | 411 | 10.3 | 350 | 85.2 | 10.1 | 10 | 2.4 | 13.2 | 51 | 12.4 | 11.4 | 61 | 14.8 | 11.7 |
| 4 symptoms | 256 | 6.4 | 201 | 78.5 | 5.8 | 8 | 3.1 | 10.5 | 47 | 18.4 | 10.5 | 55 | 21.5 | 10.5 |
| 5 or more symptoms | 542 | 13.6 | 318 | 58.7 | 9.2 | 26 | 4.8 | 34.2 | 198 | 36.5 | 44.3 | 224 | 41.3 | 42.8 |
| **Fever and anosmia/dysgeusia** |  |  |  |  |  |  |  |  |  |  |  |  |  |  |
| No | 3285 | 82.4 | 3096 | 94.2 | 89.4 | 39 | 1.2 | 51.3 | 150 | 4.6 | 33.6 | 189 | 5.8 | 36.1 |

Table 5 Association of symptoms with IgG positivity
|  | IgG results |  |  |  |  |  |  |  |  |  |  |  |  |  |
| --- | --- | --- | --- | --- | --- | --- | --- | --- | --- | --- | --- | --- | --- | --- |
|  | Total |  | Negative (<12AU/mL) |  |  | Equivocal (12÷15AU/mL) |  |  | Truly Positive (>15AU/mL) |  |  | Positive (≥12AU/mL) |  |  |
|  | N | % of total | N | % of level | % of total | N | % of level | % of total | N | % of level | % of total | N | % of level | % of total |
| Anosmia/dysgeusia | 203 | 5.1 | 83 | 40.9 | 2.4 | 17 | 8.4 | 22.4 | 103 | 50.7 | 23.0 | 120 | 59.1 | 22.9 |
| Fever | 332 | 8.3 | 252 | 75.9 | 7.3 | 7 | 2.1 | 9.2 | 73 | 22.0 | 16.3 | 80 | 24.1 | 15.3 |
| Fever and anosmia/dysgeusia | 165 | 4.1 | 31 | 18.8 | 0.9 | 13 | 7.9 | 17.1 | 121 | 73.3 | 27.1 | 134 | 81.2 | 25.6 |
| **Total** | 3985 | 100.0 | 3462 | 86.9 | 100.0 | 76 | 1.9 | 100.0 | 447 | 11.2 | 100.0 | 523 | 13.1 | 100.0 |

These results indicate that there are symptoms that best characterize the paucisymptomatic COVID-19 population and that when individuals present with fever and anosmia/dysgeusia they are likely to have SARS-CoV-2 infection.

Comorbidities (in particular cardiovascular diseases and metabolic syndrome) have been associated to a worse prognosis of COVID-19 patients ^14^. Hence we wanted to assess whether the presence of comorbidities correlated with a different incidence of IgG positivity. However, when we analyzed if there was any correlation between IgG positivity and comorbidities, we could not detect any (Supplementary Tables S4a and S4b). The same was true for the number or type of vaccination (flu, pneumococcus, tuberculosis or other vaccines) (Supplementary Tables S5a and S5b).

### IgG plasma levels and population characteristics

The frequency of IgG positive (≥12 AU/mL) individuals clearly reflected the increased exposure to the virus across the analyzed geographical areas, correlated with COVID-19-related symptoms and showed a higher proportion of positivity within young females and non-smoker subjects. However, an advantage of using a quantitative assay for IgG testing is that it is also possible to assess the magnitude of the immune response. Hence, we analyzed the data also in relation to IgG plasma levels. First, we assessed whether the level of plasma IgG correlated with positivity to the rinopharyngeal swab. To take into account a possible temporal confounding factor of symptoms detection and serological test, we compared the IgG plasma levels of swab positive or swab negative individuals with that of the ascertained COVID-19 population (between the months of March and April). We could not detect any statistically significant difference among IgG ≥12 subjects with rinopharyngeal swab positive or negative and the COVID-19 populations, suggesting that the positivity to the swab does not correlate with a higher IgG plasma level (Fig. 4a). However, we observed that physicians (including anesthesiologists and physiotherapists) had a higher level of plasmatic IgG as confirmed by statistically different odds ratios (Supplementary Table S6, *p*=0.02 and odds ratios in Supplementary Fig. S6).

**Figure 4.**
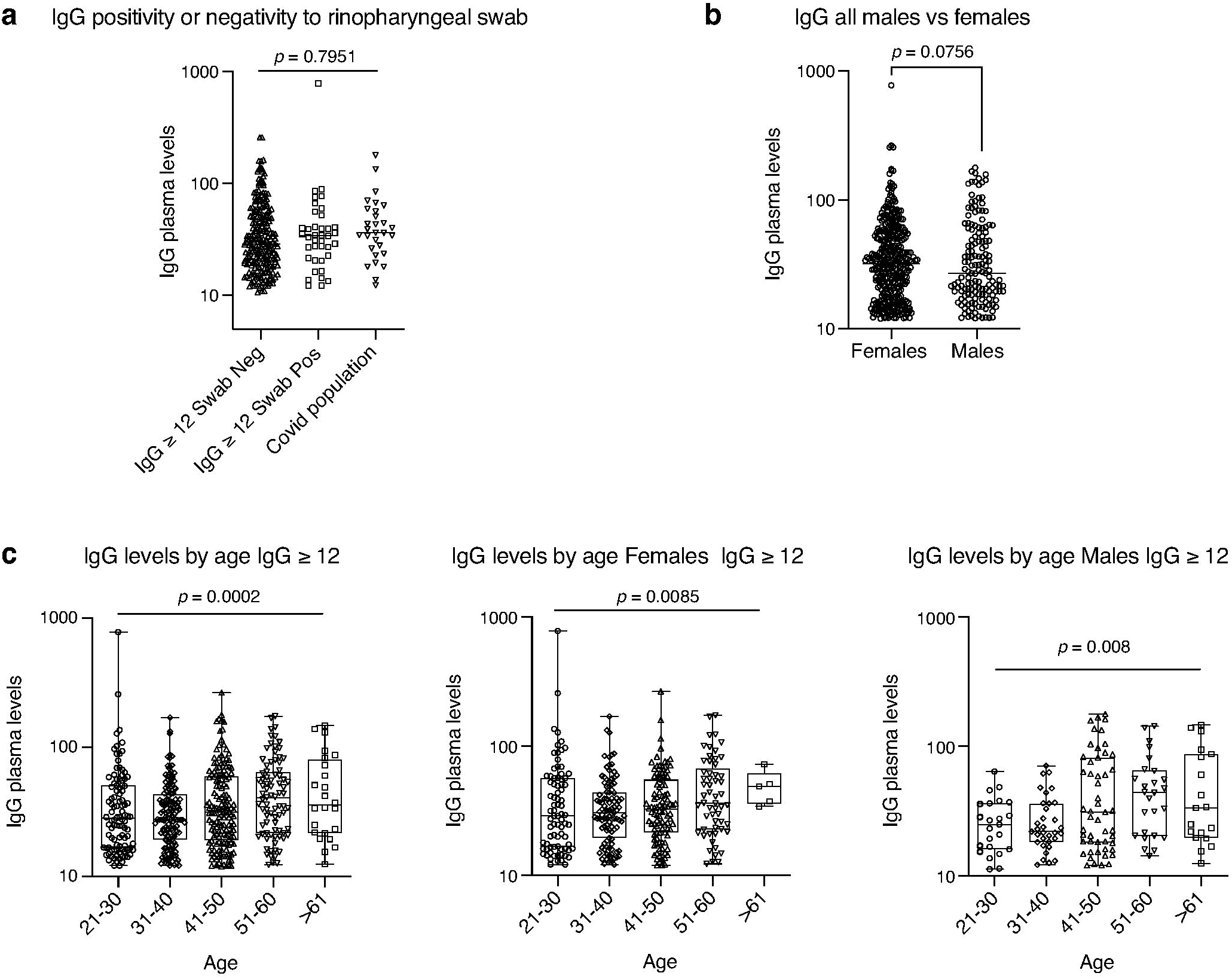
Distribution of IgG plasma levels in the different populations. a, Distribution of IgG plasma levels in the cohort of IgG positive individuals (IgG ≥ 12 AU/mL) negative or positive for the rinopharyngeal swab or ascertained COVID-19 disease. b, Distribution of IgG plasma levels in IgG positive individuals (IgG ≥ 12 AU/mL) divided by sex. c, Distribution of IgG plasma levels in IgG positive individuals (IgG ≥ 12 AU/mL) divided by age ranges and sex. In a and b *p*-values were determined using Cochran Mantel-Haenzel test for trend. In c, Cuzick’s test for trend was used: *p* = 0.0002 (total); *p* = 0.0085 (female); *p* = 0.008 (male); *p* = 0.4192 (interaction).

### IgG plasma levels BMI, gender and smoking

As we observed a higher proportion of positivity (IgG≥12 AU/mL) within females that may indicate either a higher exposure to the virus, a higher incidence of infection or a higher ability to mount an immune response, we evaluated whether this difference was paralleled by increased IgG plasma levels. We found out that there was no difference between IgG plasma levels of males versus females, suggesting a similar magnitude of the immune response (Fig. 4b, Supplementary Table S6). However, when assessing a difference of IgG plasma levels across age ranges, there was a trend towards an increased level of plasma IgG towards the older age (*p* = 0.0002, total; *p* = 0.0085, females; *p* = 0.008, males) (Fig. 4c). The difference of IgG response between females and males in relation to age remains quite intriguing and we still have to understand its relevance with the higher incidence of COVID-19 in males ^15^. Interestingly, while smoking seemed to inversely correlate with the frequency of IgG positive individuals, it did not have any effect on IgG plasma levels (Supplementary Fig. S7a) even in relation to the number of smoked cigarettes per day (Supplementary Fig. S7b and S6). On the contrary, BMI, which was not influencing the frequency of IgG positive individuals, was affecting the plasma concentration of IgG: the higher the BMI, the higher the level of IgG (Supplementary Table S6, *p*= 0.0009)

### IgG plasma levels and symptoms

As we have shown that the distribution of the IgG positive (≥12 AU/mL) population followed a sigmoidal curve with a constant level of individuals up to 7 concomitant symptoms, we thus evaluated whether there was also a correlation between the plasma level of IgG and the number of symptoms. The distribution of IgG levels in the population versus the cumulative symptoms was very similar when analyzing the whole population or those of ICH and Gavazzeni which had different proportions of IgG positive individuals (Supplementary Fig. S8). They were characterized by similar areas under the curve (All: 571; ICH: 550; Gavazzeni: 522) and the respective Receiver Operating Characteristics (ROC) curves were perfect (100%), confirming maximal specificity and sensitivity of the IgG test at IgG ≥12 AU/mL (Supplementary Fig. S8). Interestingly, we observed a direct correlation between the number of concomitant symptoms and an increase in the level of plasma IgG (Fig. 5a, Supplementary Table S7; CI are reported in the table for the different symptoms and combinations thereof, *p*<0.014). By contrast, the level of IgG in the population with values < 12 AU/mL was constant, regardless of the number of symptoms (Fig. 5b). When we analyzed the levels of IgG in relation to symptoms, we found that those subjects reporting fever, anosmia/dysgeusia, or pneumonia had all significant odds ratios (Fig. 5c). In addition, subjects with fever, cough, muscular pain, asthenia, anosmia/dysgeusia, dyspnea, chest pain, tachycardia or pneumonia, they all had higher IgG levels than those without symptoms (Supplementary Table S7, *p* values and CI are reported in the table for the different symptoms). This confirms that presence of symptoms correlates with higher IgG levels as shown in Fig. 5a. Further, combination of fever with anosmia/dysgeusia and/or dyspnea characterized populations with higher IgG plasma levels (Supplementary Table S7, *p*<0.0001, CI are reported in the table for the different symptoms and combinations thereof), also when considering the whole population regardless of IgG positivity (Fig. 5d, *p*<0.0001 in a LR test for global null hypothesis ordinal logistic analysis). When considering the level of IgG≥12 the distribution of IgG plasma levels was very similar in subjects with these symptoms (Fig. 5e), however we found statistically significant differences in odds ratios when combining fever with anosmia/dysgeusia and fever with anosmia/dysgeusia and dyspnea (Fig. 5f, *p*=0.0004 in a LR test for global null hypothesis ordinal logistic analysis). This confirms that these symptoms are the ones best characterizing SARS-CoV-2 infection. No statistically significant differences in IgG plasma levels were observed in relation to comorbidities. In steatosis/cirrhosis we had only two cases in which we found an inverse correlation but this should be confirmed on a higher number of individuals (*p*=0.0211, CI 12.30-12.70, Supplementary Table S8). Moreover, we observed an interesting inverse correlation between pneumococcal vaccination and IgG plasma levels (*p*=0.03, CI 15-29.40, Supplementary Table S9).

**Figure 5.**
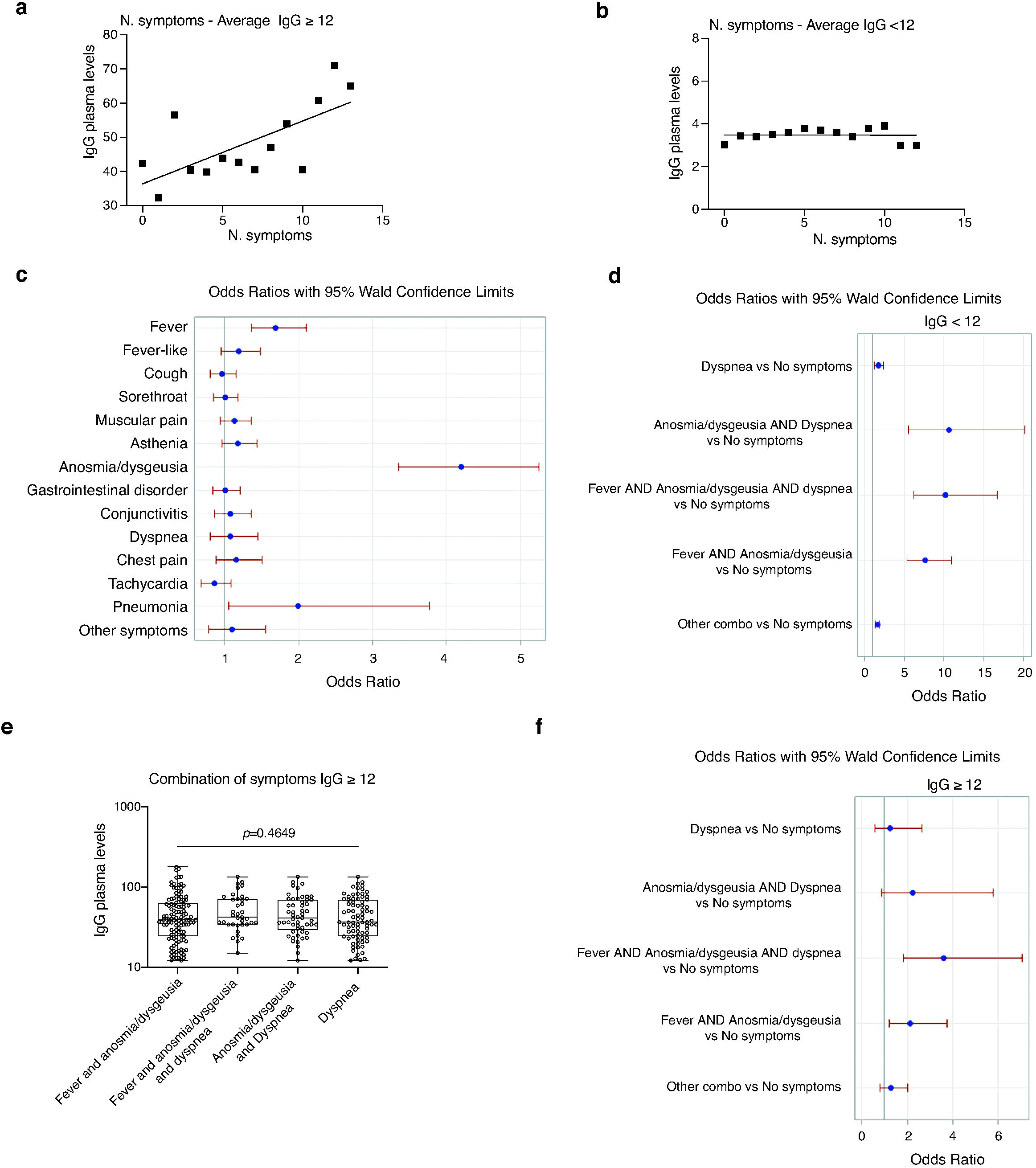
Distribution of IgG plasma levels in relation to symptoms. a, Distribution of IgG plasma levels versus the number of self-reported symptoms in IgG positive individuals (IgG ≥ 12 AU/mL). b, Distribution of IgG plasma levels versus the number of self-reported symptoms in IgG negative individuals (IgG < 12 AU/mL). c, Association between symptoms and IgG plasma levels. *p*<0.0001, LR test for global null hypothesis. Odds ratio (OR) calculated with logistic regression for ordinal data. d, Analysis of plasma levels of the whole population in relation to a combination of symptoms, *p*<0.0001. Odds ratio (OR) calculated with logistic analysis for ordinal data. e, Distribution of IgG plasma levels versus selected symptoms (alone or in combination) in individuals with IgG ≥12 AU/mL. *p*-values evaluated using Kruskal–Wallis test with Dunn’s post-hoc test. f, Analysis of plasma levels of positive subjects (IgG≥12) in relation to a combination of symptoms, *p*=0.0004. Odds ratio (OR) calculated with logistic analysis for ordinal data.

## Discussion

Here we report a comprehensive analysis of nearly 4000 individuals from different healthcare facilities representative of dramatically different levels of SARS-CoV-2 exposure in Lombardy, the most affected region by COVID-19 in Italy. We observed a range of positivity which strongly correlated with the geographical area of viral exposure from 3% in the Varese area to 43% in the Bergamo area, which were respectively the most or less COVID-affected Lombardy provinces. The proportion of IgG positive females was higher than that of males in all of the analyzed sites. However, we found a lower proportion of IgG positive individuals in females older than 60 years old than in age-matched males. This is quite intriguing as males and females are equally affected by COVID-19, but males have a worse prognosis ^16^. Through the use of a quantitative antibody test of IgG we were also able to assess the magnitude of the immune response. We found that younger males (below 40 yo) displayed reduced IgG plasma levels than older males (from 41 yo onwards). This is in line with a recent report in COVID-19 patients showing that younger patients developed lower titers of IgG ^17^ and another one showing that higher IgG titers in the plasma of convalescent individuals were associated to older age and male sex ^18^. Thus, also in the healthy population, younger males exposed to the virus develop a reduced antibody response. Hence, it is very important when analyzing the serology to SARS-CoV-2 to take into account both age and sex.

Our study differs from the one reported by Sood and colleagues in Los Angeles County^19^ as we used a quantitative antibody test and analyzed a large hospital population which ranged from healthcare professionals, researchers and administrative staff from 7 different facilities. Indirectly, we show that it is rather the environment than the hospital professional exposure which dictates the probability of contracting SARS-CoV-2 infection. Indeed, we show a higher percentage of IgG positive individuals in areas with a higher incidence of COVID-19 and among subjects who had been in contact with COVID-19 affected relatives. Further, we observed a similar proportion of IgG positive individuals among healthcare professionals functioning at hospital and administrative staff working from home. However, the magnitude of the antibody response in terms of plasma levels of IgG was higher in healthcare professionals suggesting that these individuals developed a more sustained immune response. We were also able to pinpoint 11.9% of IgG positive individuals which were completely asymptomatic and another 23.1% of paucisymptomatic subjects with 1 or 2 symptoms. This percentage differs from that observed in two recent studies analyzing SARS-CoV-2 infection via viral testing across different population types ^20,21^. Among healthcare workers, 57% of the subjects were asymptomatic/paucisymptomatic ^20^ while in a wider population in the Italian municipality of Vo’ Euganeo, that underwent lockdown soon after detecting the first case of COVID-19, the percentage of asymptomatics was 42.5 ^21^. This suggests that there is still a good proportion of individuals that probably do not develop antibodies, or develop them at low/undetectable levels. This would be in line with recent studies showing that IgG positivity is short-lived ^22^ and relatives of COVID-19 patients carry T cells specific for SARS-CoV-2 but do not seroconvert, suggesting that also IgG positivity may not detect all individuals exposed to the virus ^23^. However, if we expand our analysis to healthcare workers without symptoms but with IgG >3.8 AU/mL, and not simply those IgG ≥ 12 AU/mL, which are considered ‘positive’ by the manufacturer of the test, the percentage of positivity raises to 40%. This may indicate that there may be different exposures to the virus which result in a wide range of levels of antibody production, including those between 3.8 and 12 AU/mL. As in 283 subjects with IgG 3.8-12 AU/mL we performed a viral test, and it was negative, this indicates that they probably were exposed to the virus earlier and succeeded in eliminating it without a strong antibody response. Thus, it would be interesting to evaluate which serology test was used and which cut-off value was considered for determining IgG positivity in studies where IgG responses were negative, but T cell responses were present ^23^. In addition, some of SARS-CoV-2 viral peptides activated T cells also from unexposed individuals suggesting that some cross-reactivity could occur, maybe in response to previous infections with common cold coronaviruses ^24^.

Among the symptoms, those that characterized most the IgG positive population were fever and anosmia/dysgeusia. 81.2% of individuals presenting both anosmia/dysgeusia and fever resulted SARS-CoV-2 infected, indicating that these symptoms are strongly associated to COVID-19.

Selected vaccines such as BCG have been suggested to increase pathogen-agnostic off-target resistance to infectious agents ^25^. However, a recent report showed no differences in incidence of COVID-19 in BCG vaccinated versus non vaccinated patient population ^26^. In line with this, we did not observe a correlation between IgG positivity and BCG vaccination.

Intriguingly, we observed an inverse correlation between induction of an antibody response and smoking habit. This may indicate either a lower incidence of SARS-CoV-2 infection in smokers, or their inability to induce an immune response to the virus. The former is more likely as a large study (114,545 individuals) in Israel recently showed that the risk of SARS-CoV-2 positive testing in smoking individuals is reduced by half ^27^. When analyzing the concentrations of IgG in plasma, we found no differences between males and females, nor in smoking habits. However, we observed that the presence of some symptoms such as fever, cough, muscular pain, astenia, anosmia/dysgeusia and pneumonia correlated with higher levels of IgG than in subjects without these symptoms. In addition, we found that the presence of comorbidities did not affect IgG plasma levels except for cirrhosis. Subjects with cirrhosis had an inverse correlation with IgGs but the number of subjects was very low and this finding needs to be confirmed. The significance of this is unknown, but the finding that cirrhotic patients have higher risk of severe COVID ^28^ may suggest that this condition may affect the correct development of antibodies. Finally, we found that although very few subjects had anti-pneumoccocal vaccination, these had a reduced level of IgG, while there was no difference among those vaccinated with flu vaccine. A recent study has shown an inverse correlation between nasopharyngeal swab positivity and flu and pneumococcus vaccinations suggesting protection in vaccinated individuals ^29^. Another report on the Italian population, based on regional and not on individual data, has shown an inverse correlation between COVID-19 cases and coverage of flu vaccinations, suggesting again a protective role of flu vaccination ^30^. It is possible that flu vaccination, by inhibiting concomitant infection, may result in a better prognosis of the SARS-CoV-2 infected individuals which becomes evident only when analyzing COVID-19 cases and not asymptomatic or paucisymptomatic individuals. Why individuals vaccinated against pneumococcus should produce less antibodies remains to be evaluated.

In conclusion, our study has some limitations, as it is monitoring healthcare workers, and thus it does not reflect a comprehensive population as a good proportion of professionals are women and there is an underrepresentation of the elderly due to working age limit. However, we show that antibody testing can identify the individuals that were exposed to SARS-CoV-2 and is a powerful tool to retrospectively evaluate viral diffusion, even in asymptomatic individuals. Another limitation of the study is that we may have missed the population of subjects which do not develop an antibody response or that is reduced over time. As our study is ongoing we will assess the evolution of the IgG response over a planned follow up of one year, allowing for measuring the duration of the antibody response. Should a second wave of SARS-CoV-2 infection occur, the wide range of IgG serology in the different sites will be particularly valuable as it will allow us to assess the role of antibodies in viral protection. As we also collected the PBMC from a good proportion of our cohort, this will allow us to evaluate whether the T cell response lasts longer than the B cell response and may highlight individuals exposed to the virus which have not seroconverted, or whose level of antibodies was below 12 AU/mL. The results presented here suggest that hospital health care professionals, researchers and administrative staff can provide invaluable information to assess variables affecting the immune response to SARS-CoV-2 as a snapshot and during the follow-up.

## Methods

### Clinical Study

The study has been approved by the international review board of Istituto Clinico Humanitas for all participating institutes (clinicaltrial.gov NCT04387929). Accrual was on a voluntary basis: it started on April 28th and more than 65% of employees participated as of May 16th, 2020. 7 different centers participated to the study: Istituto Clinico Humanitas (ICH), Rozzano, Mi; Humanitas Gavazzeni, Bergamo; Humanitas Castelli, Bergamo; Humanitas Mater Domini (HMD), Castellanza (Va); Humanitas Medical Center, HMC, Varese, Mi; Humanitas University, Pieve Emanuele (Mi); Humanitas San Pio X, Milano. All participants signed an informed consent and filled a questionnaire (Supplementary Data S1) before blood collection. In order to be tested, subjects had to fill in the questionnaire. Only after filling the questionnaire in its entirety, would the individuals be scheduled for blood sampling. Hence, we obtained a 100% complete data.

All subjects with IgG > 12 AU/mL underwent a rinopharingeal swab for SARS-CoV-2 viral RNA detection.

### SARS-CoV-2 RT-PCR

Rinopharyngeal swab were tested with a commercial RT-PCR assay (AllplexTM2019-nCoV Assay – Seegene, Seoul, South Korea), according to manufacturer’s instruction. RNA extraction was performed using Seegene Nimbus, a liquid handler workstation, Real-time PCR was run on a CFX96 TMDx thermocycler (Bio-Rad Laboratories, Inc, CA, USA) and subsequently interpreted by Seegene’s Viewer software. The test target three viral genes (*E, RdRp* and *N*).

### IgG determination

For the determination of IgG anti SARS-CoV-2 the Liaison SARS-CoV-2 S1/S2 IgG assay (DiaSorin, Saluggia (VC), Italy) was used ^31^. The method is an indirect chemiluminescence immunoassay for the determination of anti-S1 and anti-S2 specific antibodies. Intra- and inter-assay coefficient of variation are <1.9% and <3.7% respectively. In the datasheet of the test, in a cohort of 304 COVID-19 patients 97.5% of the samples above 15 AU/mL and 86% of those with IgG>12 AU/mL resulted positive for neutralizing antibodies with a titer >1:40 in the Plaque Reduction Neutralization Test (PRNT).

The sensitivity of the test as reported by the manufacturer is 90.4% (79.4% - 95.8%) while the specificity is 98.5% (97.5% - 99.2%).

### Statistics

This was a cross sectional study aimed at determining the frequency of plasma anti-SARS-CoV-2 IgG antibodies in nearly 4000 (3985) employees of 7 different healthcare facilities located in Lombardy.

The serological determination was offered to all the employees of the involved sites and the anticipated refusal rate was assumed to be 10-15%. The overall IgG positivity was assumed to be around 10-15% (between 400-600 subjects).

Primary endpoint was the number of test positive subjects. Given the study size, the study was able to estimate the overall positivity with a width of 95% confidence interval equal to 2% and the positivity for subgroups of at least 200 patients with a width of 95% confidence interval equal to 9%. No power analysis was performed to calculate the sample size.

As secondary endpoints we aimed to investigate the factors associated to plasma anti-SARS-CoV-2 IgG positivity.

Basic demographic characteristics, including age, sex, smoking status, comorbidities, previous vaccinations and symptoms were recorded and their association with IgG positivity was tabulated in contingency tables and described by means of relative and absolute frequencies.

The association between these characteristics and IgG positivity was assessed by univariate logistic model and measured with the odds ratio (OR) and 95% Confidence Interval (CI) for each factor. A multivariable analysis, was also performed in order to assess the effect of characteristics of interest after adjustment for the other variables.

Furthermore, since our data have clustered structures as patients were clustered within hospitals, a multilevel model was also used to analyse data. The binary outcome (positive vs. negative serological assessment) was chosen and analysis was approached with a generalized linear model with the logic link, which is the logistic regression model. In our multilevel analysis there was a patient level and an hospital level.

We produced two models. In the first one we want to assess the effect of role, gender, age, BMI and cigarette smoking on the risk of positive serology. We estimated a random intercept model for hospital profiling and treat the hospital effects as random effects only. The role of subjects was considered related to the hospital attributes, while the subject level attributes were age (grouped in >60, ≤60 years) and gender, BMI (≤20, ≤25, ≤30, ≤35, >35) and number of smoked cigarettes/day (no,≤10, ≤20 >20). BMI and cigarette smoking were considered as continuous variables. The others variables are all categorical. The interaction term of age*gender was included in the model.

In the second one we explored the relationship between vaccinations, symptoms and serological positivity, after adjustment for the previous factors, still considering a hierarchical model. This was obtained adding the symptoms, defined as binary variables, to the previous models. Summary statistical measures were the odds ratio with relative confidence interval. SAS GLIMMIX procedure (SAS© 9.4 release) was used for the purpose of analysis.

The same approach was used also for analysis of plasma levels. SAS Mixed procedure (SAS© 9.4 release) was used for the purpose of analysis.

Kruskal–Wallis and non-parametric tests for trend (Cuzick’s and Mantel-Haenzel test, when appropriate) have been used for multiple comparisons using Prism 8 Graphpad. For plasma levels, odds ratio and relative 95%CI based on Wald Test presented in the figures were calculated using logistic model applied to ordinal data.

## Data Availability

Data is available upon request

## Data availability

Data supporting the findings of this study are available within the paper and its Supplementary Information files. All other data are available from the corresponding author upon reasonable requests.

## Acknowledgments

This work was partially supported by a philantropic donation by Dolce & Gabbana and by the Italian Ministry of Health (Ricerca corrente).

We would like to thank all the employees that volunteered to participate to this study, all the nurses and personnel that collected the samples and the laboratory technicians that run the serological and rinopharyngeal tests. We would also like to thank the Humanitas management and staff, Dr Patrizia Meroni, who warmly supported this study for the safety of the employees. Dr Alice Bertocchi for critical reading of the manuscript.

## Author contribution

M.T.S. coordinated the laboratory analyses and helped writing the manuscript; E.A.: coordinated the recruitment and sampling of subjects and participated in clinical study design; V.T.: performed the statistical analysis and helped writing the manuscript; S.C.: helped in analyzing the data; C.P.: contributed to data analysis and manuscript writing; M.S.: carried out the laboratory analyses; M.T.: participated in writing the clinical protocol; M.C.: coordinated the study for the Bergamo sites; A.M.: helped writing the manuscript and designing the study protocol; M.R.: conceived the study, analyzed the data and wrote the manuscript.

