## Supplementary Figures and Tables for "SARS-CoV-2 serology in 4000 health care and administrative staff across seven sites in Lombardy, Italy"

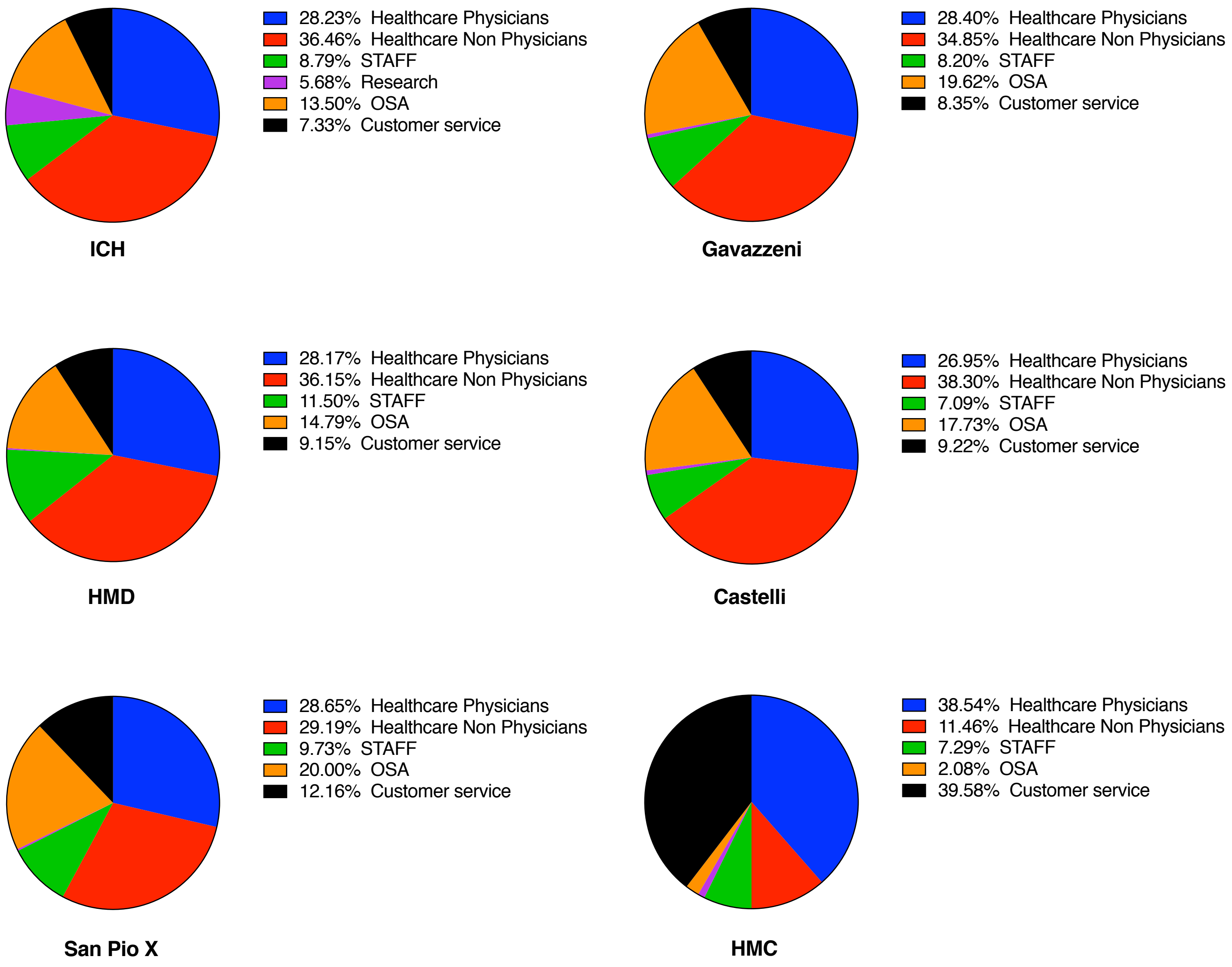

**Supplementary Figure 1. Distribution of personnel in the six hospitals analyzed.**
Pie charts show the percentage of healthcare workers participating to the study by site. Administrative staff (STAFF), nurses (OSA), customer service (check-in, admissions).

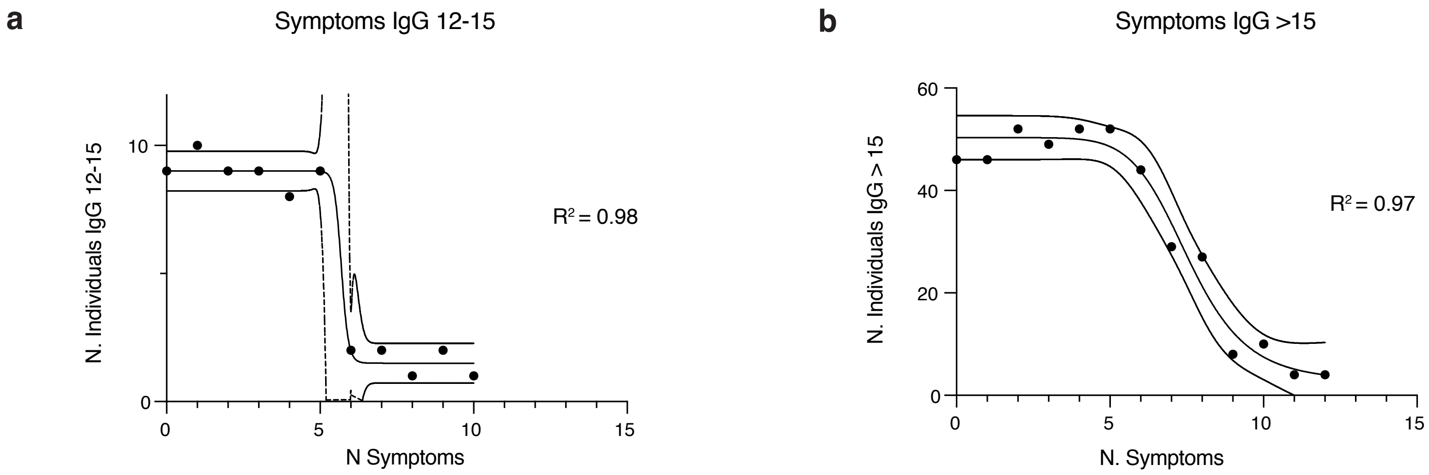

**Supplementary Figure 2. Correlation between IgG positivity and number of symptoms.**
**a**, **b**, Distribution of the IgG equivocal population (IgG 12-15 AU/mL) (**a**) and IgG truly positive population (IgG>15 AU/mL) (**b**) as number of individuals versus the number of symptoms. Both populations follow a sigmoidal, four parameter logistic curve whereby X is the number of symptoms. Distribution $R^2$ numbers are reported to demonstrate the fitness of the curve.

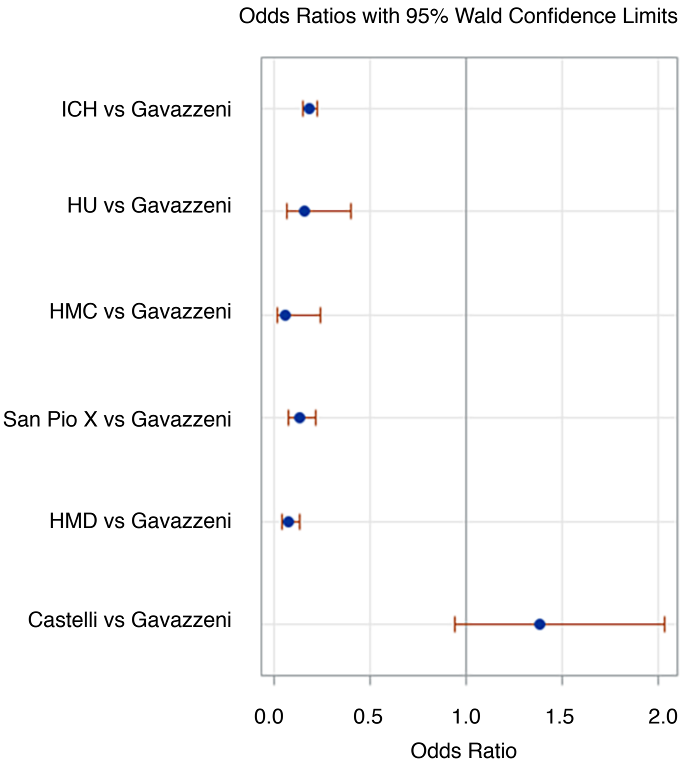

**Supplementary Figure 3. Association between site and IgG positivity.**
Odds-ratio calculated with multilevel logistic analysis.

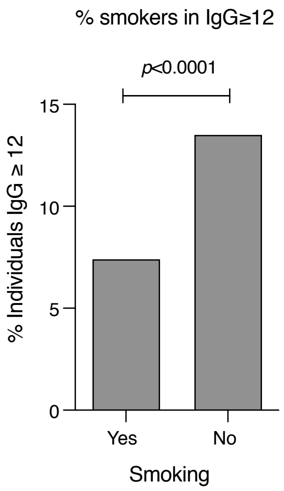

**Supplementary Figure 4. Distribution of the IgG positive population (IgG≥12 AU/mL) according to smoke.** Odds ratio calculated with multilevel logistic analysis **(**OR=0.45; 95%CI 0.34-0.60, $p$<0.0001).

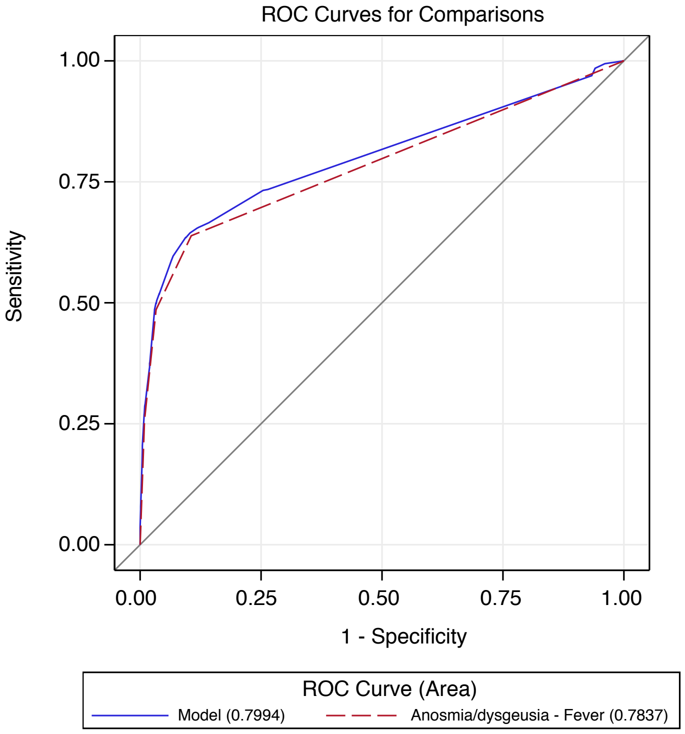

| ROC Association Statistics | | | | |
|---|---|---|---|---|
| ROC Model | Mann-Whitney | | | |
| | Area | Std Err | 95% CI | |
| Model considering all symptoms | 0.799 | 0.012 | 0.775 | 0.823 |
| Model considering only fever + anosmia/dysgeusia | 0.784 | 0.011 | 0.761 | 0.806 |

**Supplementary Figure 5. ROC analysis of relationship between IgG positivity and symptoms.** Logistic model.

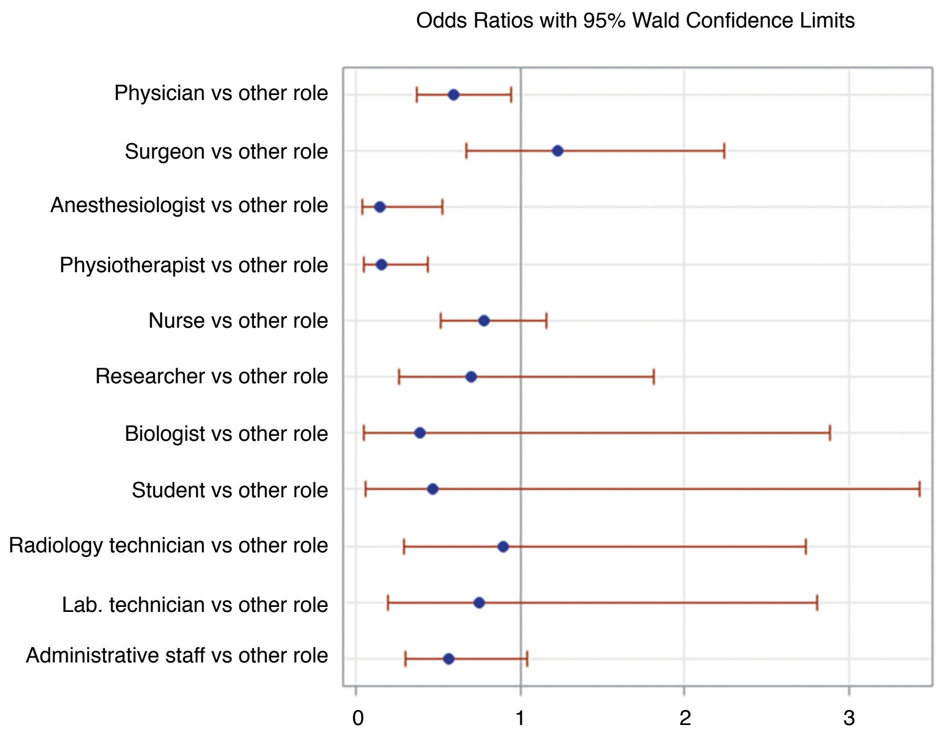

**Supplementary Figure 6. Association between role and IgG plasma levels.**
Odds-ratio calculated with logistic regression applied to ordinal data.

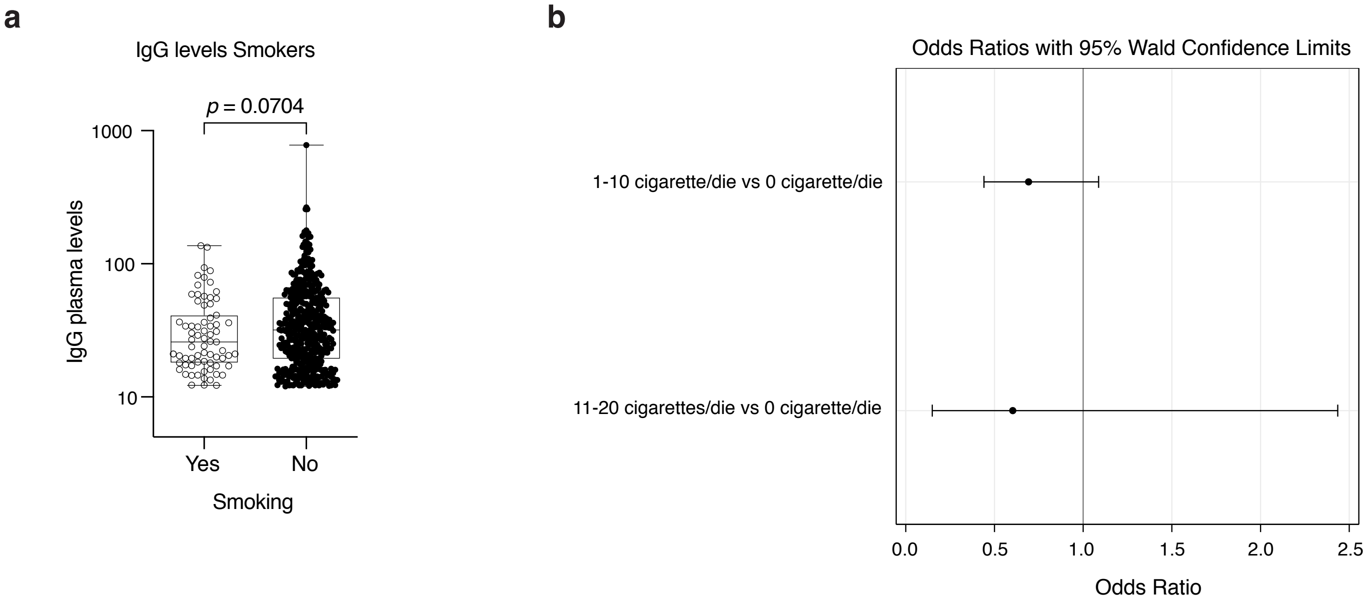

**Supplementary Figure 7. Association between smoking and IgG plasma levels.**
**a,** Distribution of the IgG positive population (IgG≥12 AU/mL) as plasma levels divided by smoking habit (yes or no). *p*-value was calculated using Kruskal-Wallis test; **b**, Odds-ratio calculated with logistic regression applied to ordinal data. LR test for global null hypothesis, p=0.207.

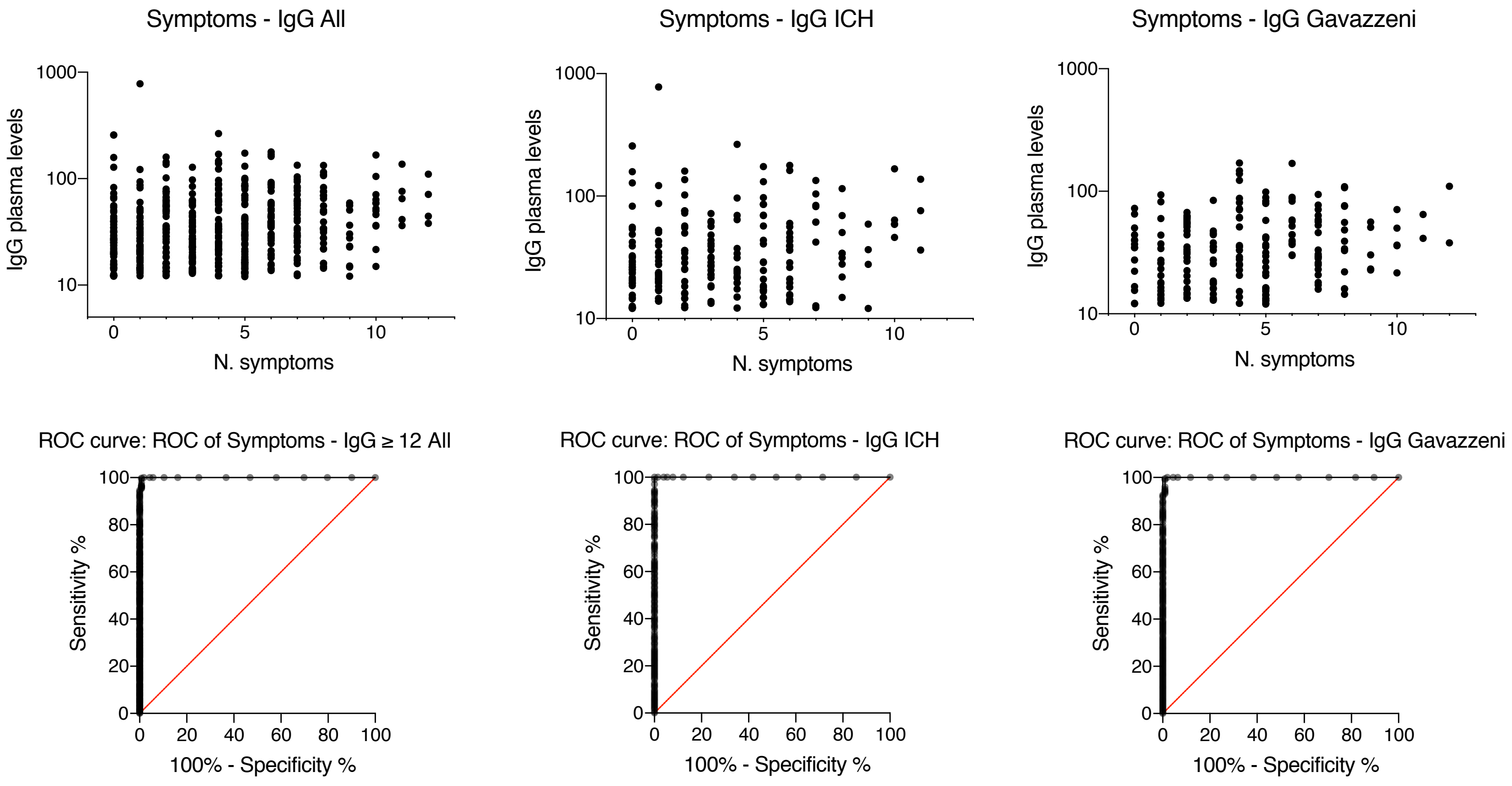

**Supplementary Figure 8. IgG plasma level distribution in the positive population (IgG ≥ 12 AU/mL) versus symptoms across all and the two major sites, ICH and Gavazzeni.** The areas under the curve are respectively: 571 All, 550 ICH, 522 Gavazzeni. Below each graph is reported the corresponding ROC curve. All of them show 100% of sensitivity and specificity.

| Supplementary Table 1 Characteristics of the population in relation to IgG positivity | | | | | | | | | | | | | | |
|---|---|---|---|---|---|---|---|---|---|---|---|---|---|---|
| | Total | | IgG results | | | | | | | | | | | |
| | | | Negative (<12AU/mL) | | | Equivocal (12÷15AU/mL) | | | Truly Positive (>15AU/mL) | | | Positive (≥12AU/mL) | | |
| | N | % of total | N | % of level | % of total | N | % of level | % of total | N | % of level | % of total | N | % of level | % of total |
| **Gender Male** | | | | | | | | | | | | | | |
| No | 2660 | 66.8 | 2288 | 86.0 | 66.1 | 57 | 2.1 | 75.0 | 315 | 11.8 | 70.5 | 372 | 14.0 | 71.1 |
| Yes | 1325 | 33.2 | 1174 | 88.6 | 33.9 | 19 | 1.4 | 25.0 | 132 | 10.0 | 29.5 | 151 | 11.4 | 28.9 |
| **Work site** | | | | | | | | | | | | | | |
| Humanitas Rozzano (ICH) | 2558 | 64.2 | 2329 | 91.0 | 67.3 | 38 | 1.5 | 50.0 | 191 | 7.5 | 42.7 | 229 | 9.0 | 43.8 |
| Humanitas University (HU) | 64 | 1.6 | 59 | 92.2 | 1.7 | 1 | 1.6 | 1.3 | 4 | 6.3 | 0.9 | 5 | 7.8 | 1.0 |
| Humanitas Medical Care (HMC) | 67 | 1.7 | 65 | 97.0 | 1.9 | . | . | . | 2 | 3.0 | 0.4 | 2 | 3.0 | 0.4 |
| Humanitas San Pio X | 250 | 6.3 | 234 | 93.6 | 6.8 | 3 | 1.2 | 3.9 | 13 | 5.2 | 2.9 | 16 | 6.4 | 3.1 |
| Humanitas Mater Domini (HMD) | 341 | 8.6 | 328 | 96.2 | 9.5 | 1 | 0.3 | 1.3 | 12 | 3.5 | 2.7 | 13 | 3.8 | 2.5 |
| Humanitas Castelli | 133 | 3.3 | 76 | 57.1 | 2.2 | 4 | 3.0 | 5.3 | 53 | 39.8 | 11.9 | 57 | 42.9 | 10.9 |
| Humanitas Gavazzeni | 572 | 14.4 | 371 | 64.9 | 10.7 | 29 | 5.1 | 38.2 | 172 | 30.1 | 38.5 | 201 | 35.1 | 38.4 |
| **Profession** | | | | | | | | | | | | | | |
| Physician | 659 | 16.5 | 569 | 86.3 | 16.4 | 22 | 3.3 | 28.9 | 68 | 10.3 | 15.2 | 90 | 13.7 | 17.2 |
| Surgeon | 287 | 7.2 | 245 | 85.4 | 7.1 | 3 | 1.0 | 3.9 | 39 | 13.6 | 8.7 | 42 | 14.6 | 8.0 |
| Anesthesiologist | 112 | 2.8 | 104 | 92.9 | 3.0 | 4 | 3.6 | 5.3 | 4 | 3.6 | 0.9 | 8 | 7.1 | 1.5 |
| Physiotherapist | 72 | 1.8 | 60 | 83.3 | 1.7 | 6 | 8.3 | 7.9 | 6 | 8.3 | 1.3 | 12 | 16.7 | 2.3 |
| Nurse | 1014 | 25.4 | 859 | 84.7 | 24.8 | 18 | 1.8 | 23.7 | 137 | 13.5 | 30.6 | 155 | 15.3 | 29.6 |
| Researcher | 123 | 3.1 | 109 | 88.6 | 3.1 | 4 | 3.3 | 5.3 | 10 | 8.1 | 2.2 | 14 | 11.4 | 2.7 |
| Biologist | 50 | 1.3 | 47 | 94.0 | 1.4 | 1 | 2.0 | 1.3 | 2 | 4.0 | 0.4 | 3 | 6.0 | 0.6 |
| Student | 28 | 0.7 | 25 | 89.3 | 0.7 | 1 | 3.6 | 1.3 | 2 | 7.1 | 0.4 | 3 | 10.7 | 0.6 |
| Radiology Technician | 94 | 2.4 | 84 | 89.4 | 2.4 | . | . | . | 10 | 10.6 | 2.2 | 10 | 10.6 | 1.9 |
| Lab. Technician | 79 | 2.0 | 72 | 91.1 | 2.1 | . | . | . | 7 | 8.9 | 1.6 | 7 | 8.9 | 1.3 |
| Administrative Staff | 315 | 7.9 | 275 | 87.3 | 7.9 | 6 | 1.9 | 7.9 | 34 | 10.8 | 7.6 | 40 | 12.7 | 7.6 |
| Other role | 1152 | 28.9 | 1013 | 87.9 | 29.3 | 11 | 1.0 | 14.5 | 128 | 11.1 | 28.6 | 139 | 12.1 | 26.6 |
| **Total** | 3985 | 100.0 | 3462 | 86.9 | 100.0 | 76 | 1.9 | 100.0 | 447 | 11.2 | 100.0 | 523 | 13.1 | 100.0 |

| Supplementary Table 2 Distribution of IgG positivity by sex across the different sites | | | | | | | |
|---|---|---|---|---|---|---|---|
| | | **Serology** | | | | **Total** | |
| | | **negative** | | **positive** | | | |
| | | **N** | **%** | **N** | **%** | **N** | **%** |
| **Working site** | | | | | | | |
| Humanitas Rozzano (ICH) | | 2329 | 91.0 | 229 | 9.0 | 2558 | 100.0 |
| Humanitas University (HU) | | 59 | 92.2 | 5 | 7.8 | 64 | 100.0 |
| Humanitas Medical Care (HMC) | | 65 | 97.0 | 2 | 3.0 | 67 | 100.0 |
| Humanitas San Pio X | | 234 | 93.6 | 16 | 6.4 | 250 | 100.0 |
| Humanitas Mater Domini (HMD) | | 328 | 96.2 | 13 | 3.8 | 341 | 100.0 |
| Humanitas Castelli | | 76 | 57.1 | 57 | 42.9 | 133 | 100.0 |
| Humanitas Gavazzeni | | 371 | 64.9 | 201 | 35.1 | 572 | 100.0 |
| Humanitas Milan | | 3015 | 91.9 | 265 | 8.1 | 3280 | 100.0 |
| **Males** | | | | | | | |
| No | | 2288 | 86.0 | 372 | 14.0 | 2660 | 100.0 |
| Yes | | 1174 | 88.6 | 151 | 11.4 | 1325 | 100.0 |
| **Working site** | **Males** | | | | | | |
| Humanitas Rozzano (ICH) | No | 1515 | 90.7 | 156 | 9.3 | 1671 | 100.0 |
| | Yes | 814 | 91.8 | 73 | 8.2 | 887 | 100.0 |
| Humanitas University (HU) | No | 37 | 90.2 | 4 | 9.8 | 41 | 100.0 |
| | Yes | 22 | 95.7 | 1 | 4.3 | 23 | 100.0 |
| Humanitas Medical Care (HMC) | No | 48 | 96.0 | 2 | 4.0 | 50 | 100.0 |
| | Yes | 17 | 100.0 | . | . | 17 | 100.0 |
| Humanitas San Pio X | No | 152 | 93.3 | 11 | 6.7 | 163 | 100.0 |
| | Yes | 82 | 94.3 | 5 | 5.7 | 87 | 100.0 |
| Humanitas Mater Domini (HMD) | No | 230 | 96.2 | 9 | 3.8 | 239 | 100.0 |
| | Yes | 98 | 96.1 | 4 | 3.9 | 102 | 100.0 |
| Humanitas Castelli | No | 56 | 57.1 | 42 | 42.9 | 98 | 100.0 |
| | Yes | 20 | 57.1 | 15 | 42.9 | 35 | 100.0 |
| Humanitas Gavazzeni | No | 250 | 62.8 | 148 | 37.2 | 398 | 100.0 |
| | Yes | 121 | 69.5 | 53 | 30.5 | 174 | 100.0 |
| Humanitas Milan | No | 1982 | 91.6 | 182 | 8.4 | 2164 | 100.0 |
| | Yes | 1033 | 92.6 | 83 | 7.4 | 1116 | 100.0 |
| **Total** | | 3462 | 86.9 | 523 | 13.1 | 3985 | 100.0 |

| Supplementary Table 3 Sensitivity, specificity and positive Likelihood ratio of symptoms | | | | | | |
|---|---|---|---|---|---|---|
| **Symptoms** | **True pos** | **False pos** | **True neg** | **False neg** | **LR pos** | **LR neg** |
| Fever | 40.9% | 8.2% | 91.8% | 59.1% | 5.01 | 1.55 |
| Low-grade Fever | 78.0% | 9.0% | 91.0% | 78.0% | 8.63 | 1.17 |
| Cough | 38.0% | 20.6% | 79.4% | 62.0% | 1.85 | 1.28 |
| Sore Throath/Runny nose | 43.4% | 30.9% | 69.1% | 56.6% | 1.40 | 1.22 |
| Muscle pain | 52.2% | 21.8% | 78.2% | 62.0% | 2.39 | 1.26 |
| Asthenia | 44.7% | 14.8% | 85.2% | 55.3% | 3.01 | 1.54 |
| Anosmia/Dysgeusia | 48.6% | 3.3% | 96.7% | 51.4% | 14.75 | 1.88 |
| Gastrointestinal disorders | 32.5% | 18.8% | 81.2% | 67.5% | 1.73 | 1.20 |
| Conjunctivitis | 16.3% | 9.2% | 90.8% | 83.7% | 1.77 | 1.08 |
| Dyspnea | 16.8% | 4.9% | 95.1% | 83.2% | 3.41 | 1.14 |
| Chest pain | 18.0% | 6.6% | 93.4% | 82.0% | 2.74 | 1.14 |
| Tachycardia | 15.7% | 10.1% | 89.9% | 84.3% | 1.55 | 1.07 |
| Pneumonia | 5.5% | 0.1% | 99.9% | 94.5% | 38.39 | 1.06 |
| Other symptoms | 6.9% | 3.5% | 96.5% | 93.1% | 1.97 | 1.04 |
| Fever & Anosmia/Dysgeusia | 25.6% | 0.9% | 99.1% | 74.4% | 28.61 | 1.33 |

| Supplementary Table 4a Correlation between comorbidities and IgG positivity | | | | | | | | | | |
|---|---|---|---|---|---|---|---|---|---|---|
| | Total | | IgG results | | | | | | | |
| | | | Negative (<12AU/mL) | | Equivocal (12÷15AU/mL) | | Truly Positive (>15AU/mL) | | Positive (≥12AU/mL) | |
| | N | % of total | N | % | N | % | N | % | N | % |
| **COPD** | | | | | | | | | | |
| No | 3968 | 99.6 | 3448 | 86.9 | 76 | 1.9 | 444 | 11.2 | 520 | 13.1 |
| Yes | 17 | 0.4 | 14 | 82.4 | . | . | 3 | 17.6 | 3 | 17.6 |
| **Asthma** | | | | | | | | | | |
| No | 3741 | 93.9 | 3249 | 86.8 | 72 | 1.9 | 420 | 11.2 | 492 | 13.2 |
| Yes | 244 | 6.1 | 213 | 87.3 | 4 | 1.6 | 27 | 11.1 | 31 | 12.7 |
| **Dyslipidemia/High Cholesterol** | | | | | | | | | | |
| No | 3619 | 90.8 | 3138 | 86.7 | 70 | 1.9 | 411 | 11.4 | 481 | 13.3 |
| Yes | 366 | 9.2 | 324 | 88.5 | 6 | 1.6 | 36 | 9.8 | 42 | 11.5 |
| **Active NPL** | | | | | | | | | | |
| No | 3981 | 99.9 | 3458 | 86.9 | 76 | 1.9 | 447 | 11.2 | 523 | 13.1 |
| Yes | 4 | 0.1 | 4 | 100.0 | . | . | . | . | . | . |
| **History of NPL** | | | | | | | | | | |
| No | 3874 | 97.2 | 3365 | 86.9 | 73 | 1.9 | 436 | 11.3 | 509 | 13.1 |
| Yes | 111 | 2.8 | 97 | 87.4 | 3 | 2.7 | 11 | 9.9 | 14 | 12.6 |
| **Chronic heart failure** | | | | | | | | | | |
| No | 3982 | 99.9 | 3459 | 86.9 | 76 | 1.9 | 447 | 11.2 | 523 | 13.1 |
| Yes | 3 | 0.1 | 3 | 100.0 | . | . | . | . | . | . |
| **Hypertension** | | | | | | | | | | |
| No | 3637 | 91.3 | 3159 | 86.9 | 71 | 2.0 | 407 | 11.2 | 478 | 13.1 |
| Yes | 348 | 8.7 | 303 | 87.1 | 5 | 1.4 | 40 | 11.5 | 45 | 12.9 |
| **History of CHD** | | | | | | | | | | |
| No | 3960 | 99.4 | 3438 | 86.8 | 76 | 1.9 | 446 | 11.3 | 522 | 13.2 |
| Yes | 25 | 0.6 | 24 | 96.0 | . | . | 1 | 4.0 | 1 | 4.0 |
| **Atrial Fibrillation** | | | | | | | | | | |
| No | 3957 | 99.3 | 3439 | 86.9 | 76 | 1.9 | 442 | 11.2 | 518 | 13.1 |
| Yes | 28 | 0.7 | 23 | 82.1 | . | . | 5 | 17.9 | 5 | 17.9 |
| **History of TIA/stroke** | | | | | | | | | | |
| No | 3973 | 99.7 | 3451 | 86.9 | 76 | 1.9 | 446 | 11.2 | 522 | 13.1 |
| Yes | 12 | 0.3 | 11 | 91.7 | . | . | 1 | 8.3 | 1 | 8.3 |
| **Steatosis/Cirrhosis** | | | | | | | | | | |
| No | 3968 | 99.6 | 3447 | 86.9 | 74 | 1.9 | 447 | 11.3 | 521 | 13.1 |
| Yes | 17 | 0.4 | 15 | 88.2 | 2 | 11.8 | . | . | 2 | 11.8 |
| **Other Hepatic diseases** | | | | | | | | | | |
| No | 3962 | 99.4 | 3442 | 86.9 | 75 | 1.9 | 445 | 11.2 | 520 | 13.1 |

| Supplementary Table 4a Correlation between comorbidities and IgG positivity | | | | | | | | | | |
|---|---|---|---|---|---|---|---|---|---|---|
| | Total | | IgG results | | | | | | | |
| | | | Negative (<12AU/mL) | | Equivocal (12÷15AU/mL) | | Truly Positive (>15AU/mL) | | Positive (≥12AU/mL) | |
| | N | % of total | N | % | N | % | N | % | N | % |
| Yes | 23 | 0.6 | 20 | 87.0 | 1 | 4.3 | 2 | 8.7 | 3 | 13.0 |
| **Chronic kidney failure** | | | | | | | | | | |
| No | 3977 | 99.8 | 3455 | 86.9 | 76 | 1.9 | 446 | 11.2 | 522 | 13.1 |
| Yes | 8 | 0.2 | 7 | 87.5 | . | . | 1 | 12.5 | 1 | 12.5 |
| **Rheumatoid Arthritis** | | | | | | | | | | |
| No | 3921 | 98.4 | 3410 | 87.0 | 72 | 1.8 | 439 | 11.2 | 511 | 13.0 |
| Yes | 64 | 1.6 | 52 | 81.3 | 4 | 6.3 | 8 | 12.5 | 12 | 18.8 |
| **Other Immune system diseases** | | | | | | | | | | |
| No | 3726 | 93.5 | 3242 | 87.0 | 73 | 2.0 | 411 | 11.0 | 484 | 13.0 |
| Yes | 259 | 6.5 | 220 | 84.9 | 3 | 1.2 | 36 | 13.9 | 39 | 15.1 |
| **Diabetes** | | | | | | | | | | |
| No | 3943 | 98.9 | 3421 | 86.8 | 76 | 1.9 | 446 | 11.3 | 522 | 13.2 |
| Yes | 42 | 1.1 | 41 | 97.6 | . | . | 1 | 2.4 | 1 | 2.4 |
| **Gout** | | | | | | | | | | |
| No | 3980 | 99.9 | 3457 | 86.9 | 76 | 1.9 | 447 | 11.2 | 523 | 13.1 |
| Yes | 5 | 0.1 | 5 | 100.0 | . | . | . | . | . | . |
| **Other comorbidities** | | | | | | | | | | |
| No | 3584 | 89.9 | 3101 | 86.5 | 72 | 2.0 | 411 | 11.5 | 483 | 13.5 |
| Yes | 401 | 10.1 | 361 | 90.0 | 4 | 1.0 | 36 | 9.0 | 40 | 10.0 |
| **Number of comorbidities** | | | | | | | | | | |
| 0 comorbidity | 2538 | 63.7 | 2192 | 86.4 | 51 | 2.0 | 295 | 11.6 | 346 | 13.6 |
| 1 comorbidity | 1062 | 26.6 | 929 | 87.5 | 18 | 1.7 | 115 | 10.8 | 133 | 12.5 |
| 2 comorbidities | 273 | 6.9 | 244 | 89.4 | 7 | 2.6 | 22 | 8.1 | 29 | 10.6 |
| 3 comorbidities | 82 | 2.1 | 71 | 86.6 | . | . | 11 | 13.4 | 11 | 13.4 |
| 4 or more comorbidities | 30 | 0.8 | 26 | 86.7 | . | . | 4 | 13.3 | 4 | 13.3 |
| **Total** | 3985 | 100.0 | 3462 | 86.9 | 76 | 1.9 | 447 | 11.2 | 523 | 13.1 |

| Supplementary Table 4b: Summary measures of association of comorbidity with IgG positivity | | | | |
|---|---|---|---|---|
| Comorbidity | Odds ratio | 95% CI | | P value |
| COPD | 3.17 | 0.79 | 12.71 | 0.1034 |
| Asthma | 0.91 | 0.59 | 1.41 | 0.6664 |
| Dyslipidemia/High Cholesterolemia | 0.94 | 0.64 | 1.39 | 0.7661 |
| Active NPL | NM | NM | NM | NM |
| History of NPL | 0.98 | 0.52 | 1.82 | 0.9398 |
| Chronic heart failure | NM | NM | NM | NM |
| Hypertension | 1.13 | 0.76 | 1.67 | 0.5401 |
| History of CHD | 0.52 | 0.06 | 4.30 | 0.5452 |
| Atrial Fibrillation | 1.29 | 0.39 | 4.26 | 0.6712 |
| History of TIA/Stroke | 0.97 | 0.11 | 8.41 | 0.9757 |
| Steatosis/ Cyrrhosis | 0.81 | 0.16 | 3.99 | 0.7972 |
| Other hepatic diseases | 0.47 | 0.10 | 2.24 | 0.3456 |
| Chronic kidney failure | 0.50 | 0.05 | 4.93 | 0.5493 |
| Rheumatoid arthritis | 1.92 | 0.91 | 4.04 | 0.0858 |
| Other Immune system diseases | 1.08 | 0.72 | 1.64 | 0.7013 |
| Diabetes | 0.19 | 0.02 | 1.42 | 0.1055 |
| Gout | NM | NM | NM | NM |
| Other comorbidities | 0.65 | 0.44 | 0.95 | 0.0274 |
| Number of comorbidities | 0.92 | 0.80 | 1.06 | 0.2414 |
| Multilevel logistic analysis, considering subjects nested in the hospital site. Adjusted for role, age (cut off = 60 years), gender, BMI, smoking habits. NM = not measurable | | | | |

| Supplementary Table 5a Correlation between IgG positivity and vaccinations | | | | | | | | | | |
|---|---|---|---|---|---|---|---|---|---|---|
| | Total | | IgG results | | | | | | | |
| | | | Negative (<12AU/mL) | | Equivocal (12÷15AU/mL) | | Truly Positive (>15AU/mL) | | Positive (≥12AU/mL) | |
| | N | % of total | N | % | N | % | N | % | N | % |
| **Recent Influenza vaccine** | | | | | | | | | | |
| No | 2774 | 69.6 | 2414 | 87.0 | 47 | 1.7 | 313 | 11.3 | 360 | 13.0 |
| Yes | 1211 | 30.4 | 1048 | 86.5 | 29 | 2.4 | 134 | 11.1 | 163 | 13.5 |
| **Anti-pneumococcal vaccine** | | | | | | | | | | |
| No | 3851 | 96.6 | 3351 | 87.0 | 68 | 1.8 | 432 | 11.2 | 500 | 13.0 |
| Yes | 134 | 3.4 | 111 | 82.8 | 8 | 6.0 | 15 | 11.2 | 23 | 17.2 |
| **Anti TBC Vaccine** | | | | | | | | | | |
| No | 3599 | 90.3 | 3122 | 86.7 | 66 | 1.8 | 411 | 11.4 | 477 | 13.3 |
| Yes | 386 | 9.7 | 340 | 88.1 | 10 | 2.6 | 36 | 9.3 | 46 | 11.9 |
| **Other vaccines** | | | | | | | | | | |
| No | 3703 | 92.9 | 3214 | 86.8 | 71 | 1.9 | 418 | 11.3 | 489 | 13.2 |
| Yes | 282 | 7.1 | 248 | 87.9 | 5 | 1.8 | 29 | 10.3 | 34 | 12.1 |
| **Number of vaccinations** | | | | | | | | | | |
| 0 vaccination | 2303 | 57.8 | 2000 | 86.8 | 37 | 1.6 | 266 | 11.6 | 303 | 13.2 |
| 1 vaccination | 1404 | 35.2 | 1224 | 87.2 | 27 | 1.9 | 153 | 10.9 | 180 | 12.8 |
| 2 vaccinations | 231 | 5.8 | 197 | 85.3 | 11 | 4.8 | 23 | 10.0 | 34 | 14.7 |
| 3 or more vaccinations | 47 | 1.2 | 41 | 87.2 | 1 | 2.1 | 5 | 10.6 | 6 | 12.8 |
| **Total** | 3985 | 100.0 | 3462 | 86.9 | 76 | 1.9 | 447 | 11.2 | 523 | 13.1 |

| Supplementary Table 5b Summary measures of association of vaccinations with IgG positivity | | | | |
|---|---|---|---|---|
| **Type of vaccination** | **Odds ratio** | **95% CI** | | ***P* value** |
| Recent Influenza vaccine | 1.15 | 0.91 | 1.44 | 0.2368 |
| Anti-pneumococcal vaccine | 1.38 | 0.81 | 2.34 | 0.2380 |
| Anti TBC Vaccine | 0.82 | 0.57 | 1.19 | 0.3029 |
| Other | 0.71 | 0.46 | 1.09 | 0.1129 |
| Multilevel logistic analysis, considering subjects nested in the hospital site. Adjusted for role, age (cut off = 60 years), gender, BMI, smoking habits. | | | | |

| Supplementary Table 6 Medians and percentiles of IgG plasma levels in relation to age, BMI, smoking, site, professional status | | | | | | | |
|---|---|---|---|---|---|---|---|
| | Positivity ≥12AU/mL | | | | | | P value* |
| | N | Median | 25°centile | 75°centile | Min | Max | |
| **Age> 60 yo** | | | | | | | 0.1212 |
| No | 497 | 30.80 | 18.50 | 53.80 | 12.10 | 778.00 | |
| Yes | 25 | 37.40 | 22.00 | 72.70 | 12.50 | 147.00 | |
| **Males** | | | | | | | 0.4793 |
| No | 371 | 32.00 | 19.20 | 53.90 | 12.10 | 778.00 | |
| Yes | 151 | 27.10 | 18.50 | 56.20 | 12.10 | 178.00 | |
| **BMI** | | | | | | | 0.0009 |
| BMI lt 20 | 64 | 28.20 | 16.00 | 53.85 | 12.80 | 128.00 | |
| BMI lt 25 | 252 | 28.70 | 17.80 | 47.95 | 12.10 | 170.00 | |
| BMI lt 30 | 127 | 34.70 | 21.80 | 59.10 | 12.10 | 169.00 | |
| BMI lt 35 | 34 | 46.20 | 19.70 | 82.10 | 12.20 | 265.00 | |
| BMI ge 35 | 9 | 38.70 | 23.90 | 62.30 | 19.70 | 778.00 | |
| **Number of smoked cigarettes** | | | | . | . | | 0.0704 |
| 0 cigarette/die | 451 | 31.90 | 19.40 | 56.10 | 12.10 | 778.00 | |
| 1-10 cigarette/die | 65 | 25.90 | 18.00 | 41.20 | 12.20 | 137.00 | |
| 11-20 cigarette/die | 6 | 23.30 | 18.00 | 34.90 | 15.50 | 61.70 | |
| 21 or more cigarette/die | 0 | . | . | . | . | . | |
| **Working site** | | | | | | | 0.2922 |
| Humanitas Rozzano (ICH) | 229 | 28.70 | 18.60 | 52.60 | 12.10 | 778.00 | |
| Humanitas University (HU) | 5 | 27.60 | 25.10 | 67.30 | 14.20 | 82.70 | |
| Humanitas Medical Care (HMC) | 2 | 20.90 | 19.20 | 22.60 | 19.20 | 22.60 | |
| Humanitas San Pio X | 16 | 43.35 | 26.25 | 73.80 | 13.40 | 128.00 | |
| Humanitas Mater Domini (HMD) | 13 | 19.00 | 16.30 | 26.70 | 14.40 | 143.00 | |
| Humanitas Castelli | 57 | 33.60 | 21.30 | 50.30 | 12.50 | 133.00 | |
| Humanitas Gavazzeni | 200 | 33.30 | 18.00 | 55.10 | 12.20 | 170.00 | |
| Humanitas Milan | 265 | 28.50 | 18.50 | 55.30 | 12.10 | 778.00 | |
| **Profession** | | | | | | | 0.0200 |
| Physician | 89 | 28.60 | 15.40 | 60.40 | 12.20 | 170.00 | |
| Surgeon | 42 | 36.25 | 21.60 | 72.10 | 12.50 | 178.00 | |
| Anesthesiologist | 8 | 15.10 | 13.50 | 37.90 | 12.80 | 76.10 | |
| Physioterapist | 12 | 16.60 | 13.40 | 21.75 | 12.20 | 64.10 | |
| Nurse | 155 | 31.80 | 20.40 | 50.90 | 12.20 | 778.00 | |
| Research | 14 | 28.20 | 15.00 | 51.90 | 12.20 | 257.00 | |
| Laboratory technician | 3 | 34.30 | 12.10 | 40.60 | 12.10 | 40.60 | |
| Student | 3 | 23.20 | 14.20 | 67.30 | 14.20 | 67.30 | |

| | | | | | | |
|---|---|---|---|---|---|---|
| Instruments technician | 17 | 31.90 | 23.80 | 48.70 | 16.20 | 63.10 |
| Staff | 17 | 31.90 | 23.80 | 48.70 | 16.20 | 63.10 |
| Other | 179 | 31.20 | 20.30 | 56.10 | 12.10 | 265.00 |
| **Total** | 522 | 31.20 | 18.60 | 54.80 | 12.10 | 778.00 |
| Kruskal Wallis test statistic for categorical variable; Cuzick's test for trend for ordinal variables. | | | | | | |

| Supplementary Table 7 Medians and percentiles of IgG plasma levels in relation to symptoms | | | | | | | |
|---|---|---|---|---|---|---|---|
| | Positivity ≥12AU/mL | | | | | | P value* |
| | N | Median | 25°centile | 75°centile | Min | Max | |
| **Fever** | | | | | | | <0.0001 |
| No | 308 | 27.10 | 17.20 | 46.50 | 12.10 | 778.00 | |
| Yes | 214 | 36.40 | 22.60 | 61.70 | 12.10 | 265.00 | |
| **Fever-like** | | | | | | | 0.0080 |
| No | 407 | 29.40 | 18.10 | 50.20 | 12.10 | 778.00 | |
| Yes | 115 | 37.70 | 22.00 | 63.10 | 12.20 | 170.00 | |
| **Cough** | | | | | | | <0.0001 |
| No | 323 | 27.10 | 16.60 | 48.70 | 12.10 | 778.00 | |
| Yes | 199 | 37.70 | 22.80 | 61.30 | 12.10 | 178.00 | |
| **Sorethroat – runny nose** | | | | | | | 0.9538 |
| No | 296 | 31.40 | 18.00 | 57.15 | 12.10 | 778.00 | |
| Yes | 226 | 30.85 | 19.50 | 50.40 | 12.10 | 170.00 | |
| **Muscular pain** | | | | | | | 0.0223 |
| No | 250 | 27.50 | 18.30 | 48.90 | 12.10 | 257.00 | |
| Yes | 272 | 33.80 | 19.35 | 58.75 | 12.10 | 778.00 | |
| **Asthenia** | | | | | | | 0.0012 |
| No | 289 | 26.90 | 18.40 | 47.50 | 12.10 | 778.00 | |
| Yes | 233 | 36.40 | 20.80 | 60.50 | 12.10 | 178.00 | |
| **Dysgeusia/Anosmia** | | | | | | | 0.0119 |
| No | 269 | 27.50 | 17.60 | 50.50 | 12.10 | 778.00 | |
| Yes | 253 | 34.30 | 21.30 | 57.20 | 12.10 | 265.00 | |
| **Gastrointestinal disorders** | | | | | | | 0.7627 |
| No | 352 | 31.20 | 19.50 | 52.30 | 12.10 | 778.00 | |
| Yes | 170 | 31.50 | 17.70 | 57.10 | 12.10 | 174.00 | |
| **Conjunctivitis** | | | | | | | 0.3557 |
| No | 438 | 30.20 | 18.40 | 55.30 | 12.10 | 778.00 | |
| Yes | 84 | 35.60 | 22.40 | 52.20 | 12.20 | 131.00 | |
| **Dyspnea** | | | | | | | 0.0044 |
| No | 434 | 29.25 | 18.00 | 50.90 | 12.10 | 778.00 | |
| Yes | 88 | 36.45 | 23.65 | 67.30 | 12.10 | 167.00 | |
| **Chest pain** | | | | | | | 0.0262 |
| No | 428 | 29.50 | 18.50 | 50.40 | 12.10 | 778.00 | |
| Yes | 94 | 36.55 | 21.50 | 63.40 | 12.20 | 265.00 | |
| **Tachycardia** | | | | | | | 0.0074 |
| No | 440 | 29.05 | 18.50 | 51.50 | 12.10 | 778.00 | |
| Yes | 82 | 39.30 | 21.50 | 69.70 | 12.20 | 178.00 | |

| Supplementary Table 7 Medians and percentiles of IgG plasma levels in relation to symptoms | | | | | | | |
|---|---|---|---|---|---|---|---|
| | Positivity ≥12AU/mL | | | | | | P value* |
| | N | Median | 25°centile | 75°centile | Min | Max | |
| **Pneumonia** | | | | | | | <0.0001 |
| No | 493 | 29.80 | 18.30 | 50.20 | 12.10 | 778.00 | |
| Yes | 29 | 76.10 | 58.80 | 105.00 | 12.10 | 178.00 | |
| **Others** | | | | | | | 0.1722 |
| No | 486 | 30.40 | 18.50 | 52.60 | 12.10 | 778.00 | |
| Yes | 36 | 39.65 | 21.75 | 58.50 | 12.10 | 174.00 | |
| **Number of symptoms** | | | | | | | 0.014 |
| 0 symptoms | 62 | 26.30 | 18.60 | 39.70 | 12.10 | 257.00 | |
| 1 symptoms | 56 | 23.25 | 17.60 | 34.50 | 12.20 | 778.00 | |
| 2 symptoms | 65 | 33.10 | 16.10 | 56.60 | 12.30 | 160.00 | |
| 3 symptoms | 61 | 26.80 | 18.00 | 45.40 | 12.80 | 128.00 | |
| 4 symptoms | 55 | 27.80 | 19.50 | 60.50 | 12.20 | 265.00 | |
| 5 or more symptoms | 223 | 36.60 | 21.60 | 61.40 | 12.10 | 178.00 | |
| **Fever and Anosmia/Dysgeusia** | | | | | | | <0.0001 |
| No | 189 | 26.10 | 16.10 | 48.70 | 12.10 | 778.00 | |
| Fever | 119 | 28.80 | 19.20 | 44.20 | 12.20 | 170.00 | |
| Anosmia/Dysgeusia | 80 | 32.95 | 21.25 | 57.85 | 12.30 | 174.00 | |
| Fever and Anosmia/Dysgeusia | 134 | 39.10 | 23.80 | 63.60 | 12.10 | 265.00 | |
| **Total** | 522 | 31.20 | 18.60 | 54.80 | 12.10 | 778.00 | |

Kruskal Wallis test statistic for categorical variable, Cuzick's test for trend for ordinal variables

| Supplementary Table 8 Medians and percentiles of IgG plasma levels in relation to comorbidities | | | | | | | |
|---|---|---|---|---|---|---|---|
| | **Positivity ≥12AU/mL** | | | | | | **P value*** |
| | **N** | **Median** | **25°centile** | **75°centile** | **Min** | **Max** | |
| **Chronic obstructive pulmonary disease** | | | | | | | 0.3286 |
| No | 519 | 31.30 | 18.50 | 55.30 | 12.10 | 778.00 | |
| Yes | 3 | 23.40 | 19.50 | 24.80 | 19.50 | 24.80 | |
| **Asthma** | | | | | | | 0.3090 |
| No | 491 | 30.60 | 18.60 | 52.70 | 12.10 | 778.00 | |
| Yes | 31 | 44.20 | 17.70 | 61.40 | 12.10 | 158.00 | |
| **Dislipidemia** | | | | | | | 0.2451 |
| No | 480 | 30.40 | 18.50 | 52.45 | 12.10 | 778.00 | |
| Yes | 42 | 36.25 | 20.40 | 65.20 | 12.30 | 178.00 | |
| **Current Neoplasia** | | | | | | | na |
| No | 522 | 31.20 | 18.60 | 54.80 | 12.10 | 778.00 | |
| Yes | 0 | . | . | . | . | . | |
| **History of Neoplasia** | | | | | | | 0.0943 |
| No | 508 | 31.40 | 19.45 | 55.05 | 12.10 | 778.00 | |
| Yes | 14 | 19.30 | 15.50 | 35.40 | 12.50 | 110.00 | |
| **Chronic cardiac failure** | | | | | | | na |
| No | 522 | 31.20 | 18.60 | 54.80 | 12.10 | 778.00 | |
| Yes | 0 | . | . | . | . | . | |
| **Hypertension** | | | | | | | 0.6716 |
| No | 477 | 31.10 | 18.50 | 54.80 | 12.10 | 778.00 | |
| Yes | 45 | 31.50 | 19.70 | 52.10 | 12.40 | 174.00 | |
| **History of Coronaropathy** | | | | | | | 0.1328 |
| No | 521 | 31.20 | 18.60 | 53.90 | 12.10 | 778.00 | |
| Yes | 1 | 98.40 | 98.40 | 98.40 | 98.40 | 98.40 | |
| **Atrial fibrillation** | | | | | | | 0.7510 |
| No | 517 | 31.20 | 18.50 | 54.80 | 12.10 | 778.00 | |
| Si | 5 | 20.40 | 20.40 | 33.10 | 19.50 | 98.40 | |
| **History of stroke** | | | | | | | 0.6328 |
| No | 521 | 31.20 | 18.60 | 54.80 | 12.10 | 778.00 | |
| Yes | 1 | 23.40 | 23.40 | 23.40 | 23.40 | 23.40 | |
| **Steatosis/Cirrhosis** | | | | | | | 0.0211 |
| No | 520 | 31.25 | 19.10 | 55.05 | 12.10 | 778.00 | |
| Yes | 2 | 12.50 | 12.30 | 12.70 | 12.30 | 12.70 | |
| **Other hepatic disorders** | | | | | | | 0.5660 |
| No | 519 | 31.20 | 18.60 | 55.30 | 12.10 | 778.00 | |
| Yes | 3 | 32.80 | 14.40 | 35.40 | 14.40 | 35.40 | |

| Supplementary Table 8 Medians and percentiles of IgG plasma levels in relation to comorbidities | | | | | | | |
|---|---|---|---|---|---|---|---|
| | Positivity ≥12AU/mL | | | | | | P value* |
| | N | Median | 25°centile | 75°centile | Min | Max | |
| **Chronic kidney failure** | | | | | | | 0.1197 |
| No | 521 | 31.20 | 18.60 | 53.90 | 12.10 | 778.00 | |
| Yes | 1 | 110.00 | 110.00 | 110.00 | 110.00 | 110.00 | |
| **Rheumatoid arthritis** | | | | | | | 0.2702 |
| No | 510 | 31.20 | 19.20 | 55.40 | 12.10 | 778.00 | |
| Yes | 12 | 31.70 | 14.60 | 41.70 | 13.90 | 56.10 | |
| **Other immune disorders** | | | | | | | 0.7754 |
| No | 483 | 31.10 | 18.50 | 55.40 | 12.10 | 778.00 | |
| Yes | 39 | 34.30 | 20.00 | 50.10 | 13.50 | 147.00 | |
| **Diabetes mellitus** | | | | | | | 0.0938 |
| No | 521 | 31.20 | 18.60 | 53.90 | 12.10 | 778.00 | |
| Yes | 1 | 167.00 | 167.00 | 167.00 | 167.00 | 167.00 | |
| **Gout** | | | | | | | na |
| No | 522 | 31.20 | 18.60 | 54.80 | 12.10 | 778.00 | |
| Yes | 0 | . | . | . | . | . | |
| **Other comorbidities** | | | | | | | 0.0082 |
| No | 482 | 29.85 | 18.40 | 52.00 | 12.10 | 778.00 | |
| Yes | 40 | 44.85 | 26.05 | 76.15 | 12.10 | 139.00 | |
| **Number of comorbidities** | | | | | | | 0.2895 |
| 0 comorbidity | 345 | 29.10 | 18.50 | 52.00 | 12.20 | 778.00 | |
| 1 comorbidity | 133 | 33.40 | 19.70 | 56.20 | 12.10 | 178.00 | |
| 2 comorbidities | 29 | 30.20 | 15.70 | 48.90 | 12.30 | 169.00 | |
| 3 comorbidities | 11 | 50.10 | 33.10 | 65.20 | 15.50 | 107.00 | |
| 4 or more comorbidities | 4 | 38.25 | 19.95 | 83.05 | 19.50 | 110.00 | |
| **Total** | 522 | 31.20 | 18.60 | 54.80 | 12.10 | 778.00 | |
| Kruskal Wallis test statistic for categorical variable, Cuzick's test for trend for ordinal variables | | | | | | | |

| Supplementary Table 9 Medians and percentiles of IgG plasma levels in relation to vaccinations | | | | | | | |
|---|---|---|---|---|---|---|---|
| | Positivity ≥12AU/mL | | | | | | P value* |
| | N | Median | 25°centile | 75°centile | Min | Max | |
| **2019/2020 Flu vaccine** | | | | | | | 0.5250 |
| No | 360 | 31.25 | 19.25 | 55.65 | 12.10 | 778.00 | |
| Yes | 162 | 30.50 | 18.50 | 51.10 | 12.20 | 160.00 | |
| **Antipneumococcus** | | | | | | | 0.0299 |
| No | 500 | 31.70 | 19.50 | 55.65 | 12.10 | 778.00 | |
| Yes | 22 | 24.70 | 15.00 | 29.40 | 12.20 | 139.00 | |
| **Anti-Tubercolar** | | | | | | | 0.6550 |
| No | 476 | 31.25 | 19.10 | 53.85 | 12.10 | 778.00 | |
| Yes | 46 | 29.25 | 18.00 | 56.30 | 12.10 | 133.00 | |
| **Other Vaccinations** | | | | | | | 0.9761 |
| No | 488 | 31.15 | 18.50 | 53.25 | 12.10 | 778.00 | |
| Yes | 34 | 32.10 | 20.30 | 58.80 | 12.20 | 96.60 | |
| **Number of vaccinations** | | | | | | | 0.5928 |
| 0 vaccination | 303 | 31.50 | 19.50 | 55.30 | 12.10 | 778.00 | |
| 1 vaccination | 180 | 30.30 | 18.55 | 53.40 | 12.10 | 160.00 | |
| 2 vaccinations | 33 | 31.60 | 14.10 | 56.60 | 12.20 | 139.00 | |
| 3 or more vaccinations | 6 | 26.65 | 18.40 | 29.40 | 13.40 | 45.40 | |
| **Total** | 522 | 31.20 | 18.60 | 54.80 | 12.10 | 778.00 | |

Kruskal Wallis test statistic for categorical variable, Cuzick's test for trend for ordinal variables

### Supplementary Data S1. Questionnaire.

#### *Questionario anamnestico di arruolamento*

##### CARATTERISTICHE SOCIO-DEMOGRAFICHE

Nome

Cognome

Data di nascita

Codice fiscale

E-mail

Genere

Peso

Altezza

Fumo sigaretta (n./die)

##### LAVORO

Dove lavori?

- Humanitas Rozzano (ICH)
- Humanitas San Pio X
- Humanitas Gavazzeni
- Humanitas Mater Domini (HMD)
- Humanitas University (HU)
- Humanitas Medical Care (HMD)

Che ruolo ha in ospedale?

- Medico
- Chirurgo
- Anestesista
- Infermiere
- OSS
- Tecnico di Radiologia
- Tecnico di Laboratorio
- Biologo
- Fisioterapista
- PARC
- Staff
- Personale Ricerca
- Studente
- Servizio Trasporto
- Servizio Pulizie
- Altro

In media nelle ultime settimane quante volte sei uscito di casa non per motivi di lavoro durante la settimana? [N.]

Negli ultimi mesi hai lavorato da casa [SI/NO]

Se sì, per quanti gg/settimana

##### ANAMNESI PATOLOGICA PROSSIMA- SINTOMATOLOGIA RIFERITA

Nel periodo dal 1° Febbraio 2020 ad oggi, ha o ha avuto uno o più dei seguenti sintomi?

- Febbre, con temperatura superiore ai 37,5°C
- Febbricola, temperatura inferiore o uguale 37,5°C per almeno tre giorni consecutivi
- Tosse
- Mal di gola e/o raffreddore
- Mal di testa
- Dolori muscolo-articolari
- Astenia
- Perdita di gusto e/o olfatto
- Disturbi gastrointestinali (diarrea, nausea, vomito)
- Congiuntivite (occhi arrossati)
- Difficoltà respiratoria/dispnea (senso di affanno a riposo)
- Dolore al petto (dolore allo sterno)
- Tachicardia/Palpitazioni
- Polmonite

##### ANAMNESI PATOLOGICA REMOTA – COMORBIDITÀ

Soffre cronicamente o le sono state diagnosticate una o più di queste condizioni?

- Broncopneumopatia Cronico Ostruttiva
- Asma
- Dislipidemia (colesterolo o trigliceridi alti) [SI/NO]
- Neoplasie attive
- Neoplasie pregresse
- Ipertensione
- Scompenso cardiaco cronico
- Coronaropatie pregresse
- Fibrillazione atriale
- Pregresso Ictus/TIA cerebrale
- Insufficienza renale cronica
- Steatosi/Cirrosi
- Altre malattie epatiche (ad es. epatite, insufficienza epatica)
- Artrite reumatoide o altre malattie reumatologiche
- Altre malattie del sistema immunitario (ad es. tiroidite, psoriasi)
- Diabete mellito
- Gotta
- Demenze

##### ALTRE CONDIZIONI DA SEGNALARE

- Interventi chirurgici nell'ultimo anno in anestesia generale

- Trapianti
- Allergie
- Gravidanza
- Immunosoppressione/ Trattamenti immunosoppressivi

#### VACCINAZIONI

Antinfluenzale nell' autunno/inverno 2019/2020 [SI/NO]
Antipneumococcica [SI/NO]
Anti-Tubercolare [SI/NO]

#### ESPOSIZIONE COVID-19

Negli ultimi 3 mesi è entrato/a in stretto contatto (contatto diretto a meno di 2 metri di distanza, o in ambiente chiuso come casa, sede di lavoro, mezzo di trasporto) con **casi accertati** o sospetti di COVID-19?

Se sì, chi era?
Collega
Paziente
Familiare
Altro_specificare
